# Tay-Sachs and Sandhoff Diseases: Diffusion tensor imaging and correlational fiber tractography findings differentiate late-onset GM2 Gangliosidosis

**DOI:** 10.1101/2024.12.13.24318793

**Authors:** Connor J. Lewis, Selby I. Chipman, Jean M. Johnston, Maria T. Acosta, Camilo Toro, Cynthia J. Tifft

## Abstract

GM2 gangliosidosis is lysosomal storage disorder caused by deficiency of the heterodimeric enzyme β-hexosaminidase A. Tay-Sachs disease is caused by variants in *HEXA* encoding the α-subunit and Sandhoff disease is caused by variants in *HEXB* encoding the β-subunit. Due to shared clinical and biochemical findings, the two have been considered indistinguishable. We applied diffusion tensor imaging (DTI) and correlational fiber tractography to assess phenotypic differences in these two diseases. 40 DTI scans from 16 Late-Onset GM2 patients (NCT00029965) with either Sandhoff (n = 4), or Tay-Sachs (n = 12) disease. DTI metrics including fractional anisotropy (FA), mean diffusivity (MD), radial diffusivity (RD), axial diffusivity (AD), and quantitative anisotropy (QA) were calculated in fiber tracts throughout the whole brain, arcuate fasciculus, corpus callosum, and cerebellum. Correlational tractography was also performed to identify fiber tracts with group wide differences in DTI metrics between Tay-Sachs and Sandhoff patients. A linear mixed effects model was used to analyze the differences between Tay-Sachs and Sandhoff patients. Tay-Sachs patients had higher MD in the left cerebellum (*p* = 0.003703), right cerebellum (*p* = 0.003435), superior cerebellar peduncle (SCP, *p* = 0.007332), and vermis (*p* = 0.01007). Sandhoff patients had higher FA in the left cerebellum (*p* = 0.005537), right cerebellum (*p* = 0.01905), SCP (*p* = 0.02844), and vermis (*p* = 0.02469). Correlational fiber tractography identified fiber tracts almost exclusively in cerebellar pathways with higher FA and QA, and lower MD, AD, and RD in Sandhoff patients compared to Tay-Sachs patients. Our study shows neurobiological differences between these two related disorders. To our knowledge, this is the first study using correlational tractography in a lysosomal storage disorder demonstrating these differences. This result indicates a greater burden of cerebellar pathology in Tay-Sachs patients compared with Sandoff patients.

## Introduction

GM2 gangliosidoses are autosomal recessive neurodegenerative lysosomal storage disorders caused by the cytotoxic accumulation of GM2 ganglioside. β-hexosaminidase A is the deficient enzyme and is responsible for degrading GM2 into GM3 as a part of the ganglioside metabolism pathway.^1,2^ Three diseases are associated with GM2 gangliosidosis, Tay-Sachs disease is characterized by biallelic variants in *HEXA* encoding the α subunit of β-hexosaminidase A. Sandhoff disease is characterized by biallelic variants in *HEXB* encoding the ß subunit resulting in deficiencies in both hexosaminidase A and B. GM2 activator deficiency, known as the AB variant, is characterized by biallelic variants in *GM2A*.^3^ The resulting accumulation of GM2 gangliosides is toxic primarily to neurons where gangliosides play a key role in central nervous system function.

The frequency of Tay-Sachs disease is estimated between 1 in 200,000 to 1 in 320,000 individuals.^4,5^ The estimated prevalence for Sandhoff disease is between 1 in 500,000 to 1 in 1,500,000 individuals.^6^ The AB activator deficiency is the rarest form of GM2 gangliosidosis with only 13 known cases reported.^6^ There are no approved therapies for GM2 gangliosidosis, however investigations into enzyme replacement therapy, substrate reduction therapy, and gene therapy as potential treatments are currently underway.^7^

Sandhoff and Tay-Sachs diseases can be further classified into three subtypes of based on symptom onset age and disease progression corresponding to residual enzyme activity.^7,8^ The infantile classifications of these diseases are the most severe with symptom onset before 6 months and death by 5 years of age.^9^ The juvenile form of these diseases is less severe with symptom onset between 2 and 6 years and death within the second decade.^10–12^ Adult or late onset GM2 gangliosidosis is the least severe form of the disease with symptom onset frequently occurring between adolescence and early adulthood, and reduced life expectancy compared to unaffected adults.^12–15^ The adult or late onset form of GM2 gangliosidosis is also differentiated from the juvenile form due to preserved cognitive function.^9^ While the infantile and juvenile forms of the activator deficiency have been described, an adult-onset form of the activator deficiency has not been described to the best of our knowledge.^16–18^

Due to similar clinical presentations, late-onset Sandhoff (LOSD) and Tay-Sachs (LOTS) have been presumed indistinguishable, however recent studies have focused on differentiating these two forms of GM2 gangliosidosis.^19,20^ Both disorders produce characteristic weakness of hip flexor, knee extensors and later on, triceps weakness. A slightly lower age of onset, psychosis, and dysarthria are more commonly associated with LOTS, whereas length dependent sensory peripheral neuropathy and burning pain in feet and hands are more typical for LOSD patients.^14,21,22^

Case studies of MRI findings in late onset GM2 gangliosidosis have demonstrated numerous findings affecting the thalamus, enlargement of the 4^th^ ventricle,^23,24^ white matter abnormalities, atrophy of the cerebral cortex,^21^ brainstem,^21^ corpus callosum,^25^ and cerebellar atrophy (including gray and white matter).^21,23,24^ Previous studies have also mentioned cerebellar atrophy and enlarged 4^th^ ventricle which are more prevalent and severe in Tay-Sachs disease as opposed to Sandhoff disease.^14,21,23,26^ Magnetic resonance spectroscopy (MRS), an imaging technique analyzing metabolite concentration *in vivo* has shown pathogenic differences associated with GM2 gangliosidosis.^27^ Furthermore, MRS has highlighted cerebellar metabolic differences between Tay-Sachs and Sandhoff disease patients.^23^

Diffusion weighted imaging (DWI) is a noninvasive neuroimaging modality with the capability of analyzing white matter microstructure based on the relative diffusion and diffusion restriction in the brain.^28^ A previous study in the GM2 mouse model of Sandhoff disease has demonstrated a reduced apparent diffusion coefficient compared with wild type mice in the cortex, striatum, and thalamus.^29^ However, we were unable to find any studies in humans.

Diffusion tensor imaging (DTI) builds on DWI by evaluating the diffusion tensor matrix, and allows for fiber tractography calculations.^30,31^ DTI metrics include fractional anisotropy (FA) which is useful in characterizing white matter integrity, mean diffusivity (MD), axial diffusivity (AD), and radial diffusivity (RD). AD is a measure of the diffusion along the principal axis or parallel along an axon where decreased AD has been associated with axonal injury.^32^ AD has been shown to increase with structural brain changes that accompany aging, Huntington’s disease and Alzheimer’s disease^33,34^ FA is a measure of directional diffusion restriction and pathogenic reductions in FA may represent reduced axonal packing, integrity.^32^ MD is an average of the diffusion and pathogenic increases in MD may represent reduced white matter integrity.^32^ RD is a measure of the diffusion perpendicular to the axon, where higher RD may represent myelin loss, axonal loss, reduced axonal packing density, or some combination of the three.^32^

Quantitative anisotropy (QA) is a newer diffusion MRI metric derived from generalized-q-sampling imaging which describes the Fourier transform between water’s diffusion and signal decay.^35^ QA utilizes the spin distribution function to calculate anisotropy and may offer improvements over traditional FA approaches in describing axonal loss.^35^ QA has shown improvements in dealing with the noise associated with DWI scans and may offer improvements in dealing with crossing fibers and complex fiber orientations, a known issue associated with FA.^36^

Correlational fiber tractography is group level analysis technique where a DTI metric is evaluated in relation to a study variable.^37^ Correlational tractography approaches have shown the potential to be more sensitive than conventional tractography approaches.^38,39^ To the best of our knowledge, this is the first study investigating sectional correlational fiber tractography to assess differences between GM2 gangliosidosis disease subtypes in brain neuronal tracts of late onset patients.

## Methods

### The Natural History of GM2 Gangliosidosis

Participants from the National Human Genome Research Institute (NHGRI) study, the “Natural History of Glycosphingolipid & Glycoprotein Storage Disorders” with a diagnosis of either LOTS or LOSD disease and at least one DWI scan were included in this analysis (NCT00029965).^40^ The NIH Institutional Review Board approved this protocol (02-HG-0107). Informed consent was completed with all patients prior to participation and all research was completed in accordance with the Declaration of Helsinki. All patient visits were conducted at the National Institutes of Health Clinical Center (Bethesda MD) between 2010 and 2020.

The distinction between LOTS and LOSD was made based on differences between β-hexosaminidase A and total β-hexosaminidase (β-hexosaminidase A + β-hexosaminidase B) activity patterns and biallelic variants in either *HEXA* or *HEXB*. Twelve LOTS and four LOSD subjects were included (Supplement A).

### DWI Acquisition and Preprocessing

40 DWI scans were acquired from 16 GM2 patients on a Phillips Achieva 3T system with an 8-channel SENSE head coil. DWI were acquired with the following parameters: TR/TE=6400/100 ms, 32-gradient encoding directions, b-values=0 and 1000 s/mm^2^, voxel size = 1.875mm×1.875mm×2.5mm, slice thickness=2.5 mm, acquisition matrix=128×128, NEX=1, FOV=24 cm. DWI were preprocessed using MRtrix3’s (MRtrix, v3.0.4)^41^ *dwifslpreproc*^42–44^ command utilizing the *dwi2mask*^45^ function followed by FSL’s (FSL, v6.0.5) *eddy*^43^ and *topup*^43,44^ functions. Preprocessed data were imported into DSI Studio (DSI Studio, v2023), where imaging was quality checked for bad slices, a U-Net mask was created, and generalized q-sampling imaging (GQI) based reconstruction was performed with a diffusion sampling length ratio of 1.25^46^

### DTI Analysis

The 40 preprocessed and reconstructed DWI scans were analyzed with a diffusion MRI connectometry in DSI Studio to map fiber tracts and calculate diffusion tensor metrics along identical pathways. An atlas based deterministic fiber tractography was performed for the whole brain, cerebellum (bilateral), inferior cerebellar peduncle (bilateral), middle cerebellar peduncle (MCP), superior cerebellar peduncle (SCP), vermis, corpus callosum, and arcuate fasciculus (bilateral) and fractional anisotropy and mean diffusivity were calculated. The angular threshold was 0 (random), the step size was 0 (random) mm. Tracks < 20 mm or > 200 mm were discarded and 1,000,000 seeds were placed.

### Correlational Tractography Analysis

A separate diffusion MRI connectometry analysis was also performed in DSI Studio (DSI Studio, v2023) to identify differences in fractional anisotropy and mean diffusivity between Sandhoff (n = 4) and Tay-Sachs (n = 12) patients. Correlational tractography was performed on the 16 preprocessed and reconstructed baseline scans of GM2 patients with a *T*-score between 2.0-4.0 for deterministic fiber tractography. A fiber tract length threshold between 10-40 voxels was also applied where fiber tracts shorter than the threshold were removed. The effects of age removed using a multiple regression model, and the false detection rate (FDR) was estimated using 4,000 random permutations and a threshold of <0.05 was applied to only include axonal loss.

### Statistical Analysis

Statistical analysis in this study was performed in R (The R Foundation, v4.3.1). Between group analysis was performed using a linear mixed effects model with age (fixed effect) as a covariate of no interest to evaluate DTI data. DTI metrics including FA, MD, RD, AD, and QA were evaluated against GM2 gangliosidosis disease subtype to determine if there was a significant difference between Tay-Sachs and Sandhoff patients. Each participant was assigned as a random effect to account for the repeated DWI scans conducted on each participant. *P*-values < 0.05 were considered significant.

## Results

### Diffusion Tensor Imaging Analysis - Fractional Anisotropy (FA)

Table I. summarizes the DTI results of fractional anisotropy (FA) between Tay-Sachs and Sandhoff patients as evaluated by linear mixed effects modeling. There was no statistical difference in FA between Tay-Sachs and Sandhoff patients throughout the whole brain (Figure 1A), corpus callosum (Figure 3A), or bilateral arcuate fasciculus (Supplement Figures B7A and B8A). There was also no statistical difference in FA in the bilateral inferior cerebellar peduncle and middle cerebellar peduncle in Sandhoff patients compared to Tay-Sachs patients. Sandhoff patients had higher FA in the bilateral cerebellum (Figure 2A, Supplement Figure B1A), superior cerebellar peduncle (Supplement Figure B5A), and vermis (Supplement Figure B6A) compared to Tay-Sachs patients.

**Table I.** Diffusion Tensor Imaging Results of FA in Atlas Fiber Tractography Pathways evaluating differences between Tay-Sachs and Sandhoff Patients.

| Pathway | Estimate | Standard Error | $\chi^2(1)$ | $p\text{-value } (>\chi^2)$ |
| --- | --- | --- | --- | --- |
| Whole Brain | 0.0070 | 0.0094 | 0.699 | 0.4032 |
| Left Cerebellum | 0.0479 | 0.0167 | 7.695 | **0.005537** |
| Right Cerebellum | 0.0391 | 0.0166 | 5.497 | **0.01905** |
| Left Inferior Cerebellar Peduncle | 0.0380 | 0.0222 | 3.103 | 0.07814 |
| Right Inferior Cerebellar Peduncle | 0.0250 | 0.0190 | 1.889 | 0.1693 |
| Middle Cerebellar Peduncle | 0.0223 | 0.0183 | 1.647 | 0.1993 |
| Superior Cerebellar Peduncle | 0.0268 | 0.0121 | 4.801 | **0.02844** |
| Vermis | 0.0498 | 0.0223 | 5.046 | **0.02469** |
| Corpus Callosum | 0.0016 | 0.0087 | 0.050 | 0.8239 |
| Left Arcuate Fasciculus | 0.0090 | 0.0104 | 0.9237 | 0.3365 |
| Right Arcuate Fasciculus | -0.0007 | 0.0124 | 0.0442 | 0.8335 |

**Figure 1.**
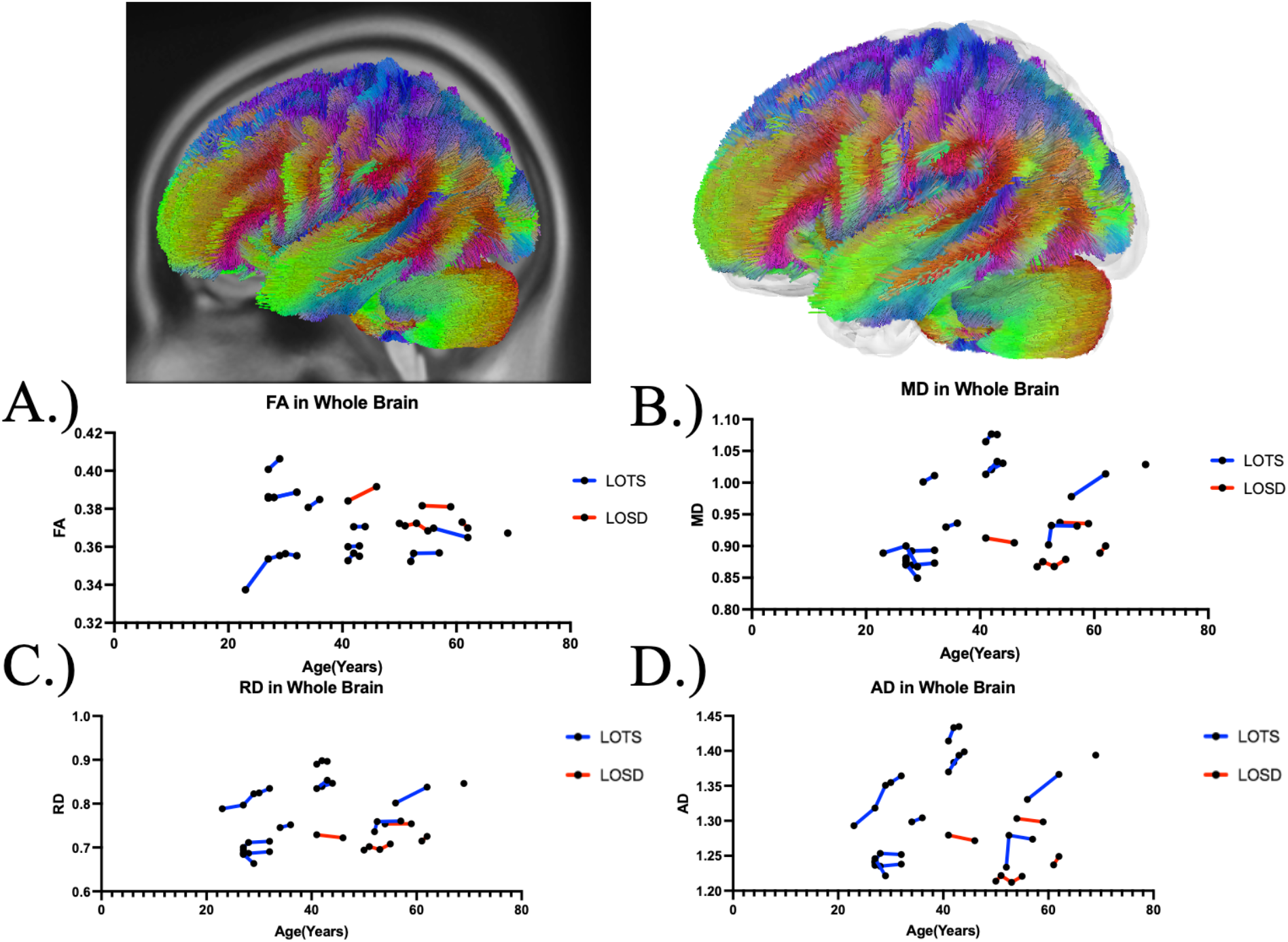
Atlas Based Fiber Tractography of the whole brain demonstrating age related effects on A.) fractional anisotropy (FA), B.) mean diffusivity (MD), C.) radial diffusivity (RD), D.) axial diffusivity (AD) between Tay-Sachs patients (blue) and Sandhoff patients (red). Tay-Sachs patients demonstrated no difference in FA (*χ*^2^(1) = 0.699, *p =* 0.005537) and increased MD (*χ*^2^(1) = 7.62, *p =* 0.005771), RD (*χ*^2^(1) = 7.28, *p =* 0.006981), and AD (*χ*^2^(1) = 7.86, *p =* 0.005066) compared to Sandhoff patients in fiber tracts throughout the whole brain when age was accounted for.

**Figure 2.**
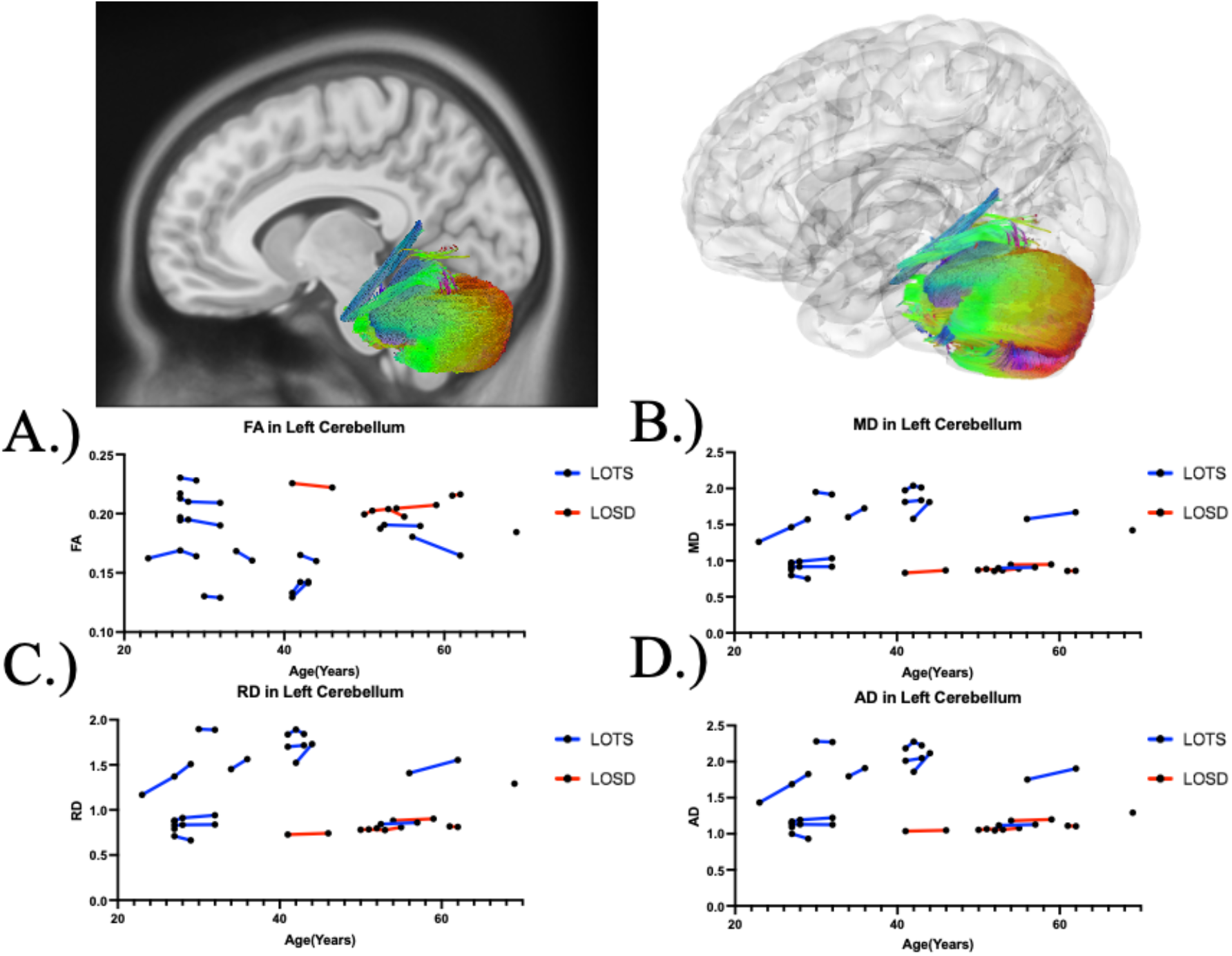
Atlas Based Fiber Tractography of the left cerebellum demonstrating age related effects on A.) fractional anisotropy (FA), B.) mean diffusivity (MD), C.) radial diffusivity (RD), D.) axial diffusivity (AD) between Tay-Sachs patients (blue) and Sandhoff patients (red). Tay-Sachs patients demonstrated decreased FA (*χ*^2^(1) = 7.695, *p =* 0.005537) and increased MD (*χ*^2^(1) = 8.42, *p =* 0.003703), RD (*χ*^2^(1) = 8.51, *p =* 0.003536), and AD (*χ*^2^(1) = 8.25, *p =* 0.004065) compared to Sandhoff patients in left cerebellar fiber tracts when age was accounted for.

**Figure 3.**
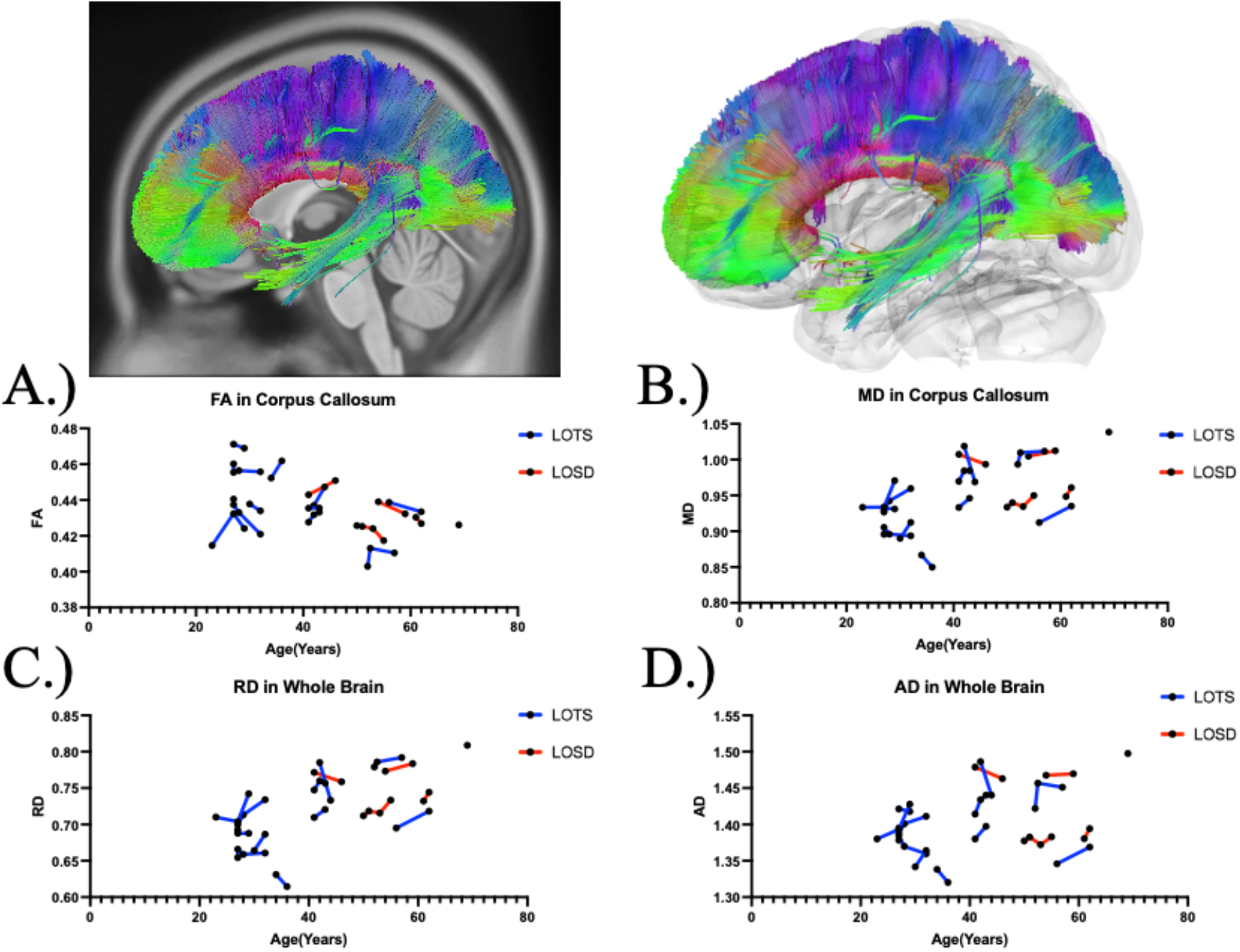
Atlas Based Fiber Tractography of the corpus callosum demonstrating age related effects on A.) fractional anisotropy (FA), B.) mean diffusivity (MD), C.) radial diffusivity (RD), D.) axial diffusivity (AD) between Tay-Sachs patients (blue) and Sandhoff patients (red). Tay-Sachs patients demonstrated decreased FA (*χ*^2^(1) = 7.695, *p =* 0.005537) and increased MD (*χ*^2^(1) = 8.42, *p =* 0.003703), RD (*χ*^2^(1) = 8.51, *p =* 0.003536), and AD (*χ*^2^(1) = 8.25, *p =* 0.004065) compared to Sandhoff patients in corpus callosum fiber tracts when age was accounted for.

### Diffusion Tensor Imaging Analysis - Mean Diffusivity (MD)

Table II. summarizes the DTI results of MD between Tay-Sachs and Sandhoff patients as evaluated by linear mixed effects modeling. MD was higher in white matter pathways throughout the whole brain (Figure 1B), bilateral cerebellum (Figure 2B and Supplement Figure B1B), bilateral inferior cerebellar peduncle (Supplement Figures B2B and B3B), MCP (Supplement Figure B4B), SCP (Supplement Figure B5B), and vermis (Supplement Figure B6B) in Tay-Sachs patients compared to Sandhoff patients. MD in the corpus callosum (Figure 3B) and bilateral arcuate fasciculus (Supplement Figures B7B and B8B) were not different between Tay-Sachs and Sandhoff patients.

**Table II.**
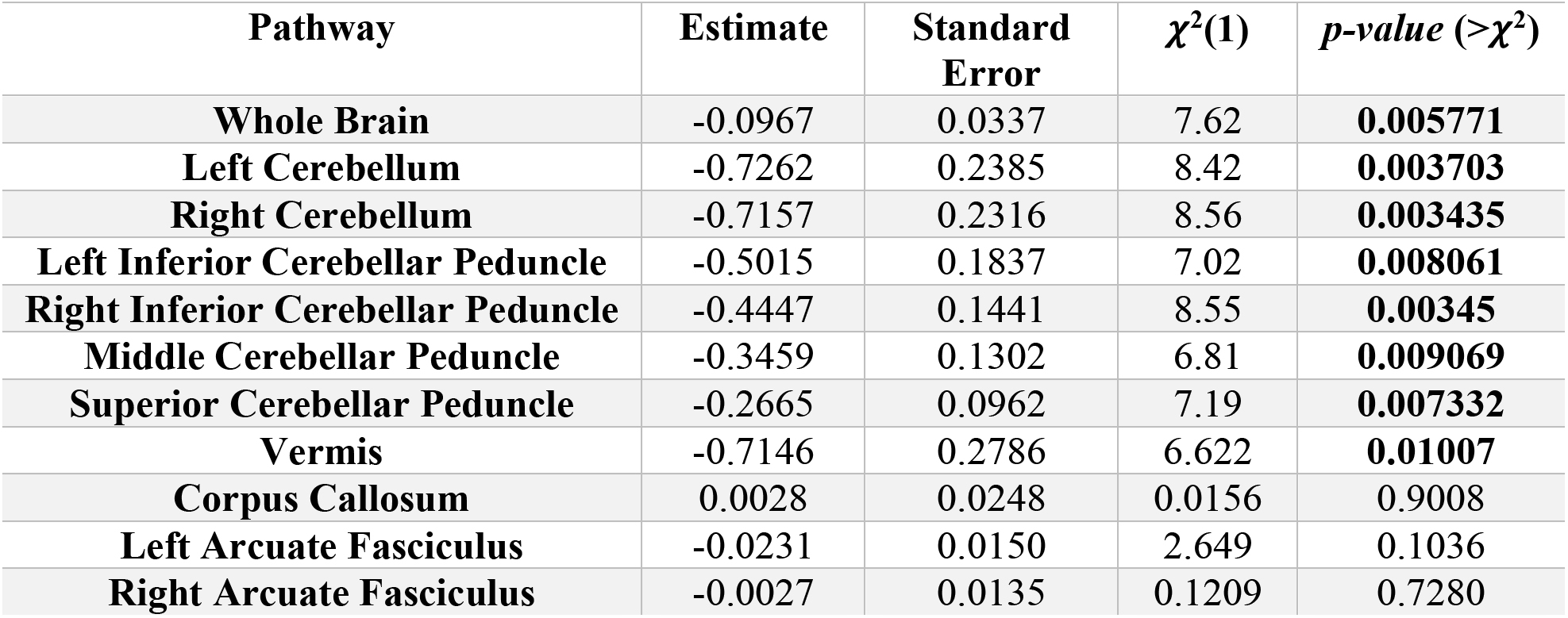
Diffusion Tensor Imaging Results of MD in Atlas Fiber Tractography Pathways evaluating differences between Tay-Sachs and Sandhoff Patients.

### Diffusion Tensor Imaging Analysis - Radial Diffusivity (RD)

Supplement Table C1 summarizes the DTI results of RD between Tay-Sachs and Sandhoff patients as evaluated by linear mixed effects modeling. RD was higher in white matter pathways throughout the whole brain (Figure 1C), bilateral cerebellum (Figure 2C and Supplement Figure B1C), bilateral inferior cerebellar peduncle (Supplement Figure B2C and B3C), MCP (Supplement Figure B4C), SCP (Supplement Figure B5C), and vermis (Supplement Figure B6C) in Tay-Sachs patients compared to Sandhoff patients. RD in the corpus callosum (Figure 3C) and bilateral arcuate fasciculus (Supplement Figures B7C and B8C) were not different between Tay-Sachs and Sandhoff patients.

### Diffusion Tensor Imaging Analysis - Axial Diffusivity (AD)

Supplement Table C2 summarizes the DTI results of AD between Tay-Sachs and Sandhoff patients as evaluated by linear mixed effects modeling. AD was higher in white matter pathways throughout the whole brain (Figure 1D), bilateral cerebellum (Figure 2D and Supplement Figure B1D), bilateral inferior cerebellar peduncle (Supplement Figures B2D and B3D), MCP (Supplement Figure B4D), SCP (Supplement Figure B5D), and vermis (Supplement Figure B6D) in Tay-Sachs patients compared to Sandhoff patients. AD in the corpus callosum (Figure 3D) and bilateral arcuate fasciculus (Supplement Figures B7D and B8D) were not different between Tay-Sachs and Sandhoff patients.

### Diffusion Tensor Imaging Analysis - Quantitative Anisotropy (QA)

Supplement Table C3 summarizes the DTI results of QA between Tay-Sachs and Sandhoff patients as evaluated by linear mixed effects modeling. QA was not statistically different between Tay-Sachs and Sandhoff patients for fiber tracts throughout the whole brain, bilateral arcuate fasciculus, bilateral cerebellum, corpus callosum, vermis, MCP, SCP, or bilateral inferior cerebellar peduncle.

### Correlational Fiber Tractography Analysis

Figures 4 and 5 show significant differences in FA and MD in neuronal fiber tracts between Sandhoff and Tay-Sachs patients as evaluated by correlational fiber tractography. Fiber tracts were identified almost exclusively in the cerebellum with higher FA and lower MD in Sandhoff patients when testing for differences based on disease subtype. No fiber tracts were evaluated to have higher FA or lower MD in Tay-Sachs patients when compared to Sandhoff patients. Higher T-score and length thresholds demonstrated less results and ultimately no results at the highest thresholds for FA and MD.

**Figure 4.**
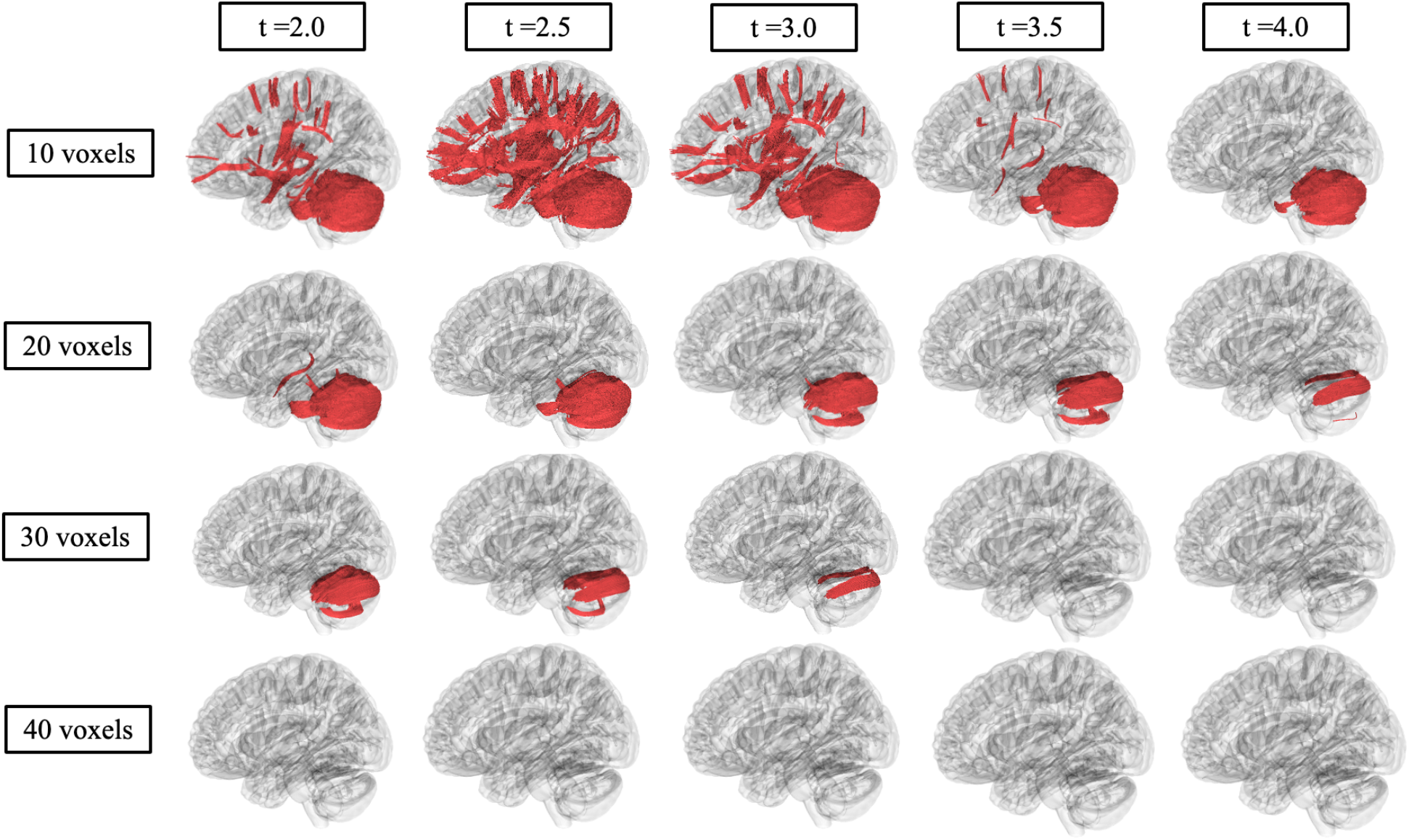
Correlational fiber tractography assessed differences in fractional anisotropy in Sandhoff and Tay-Sachs patients at varying length (voxels) and T thresholds. Fiber tracts shown in red were evaluated to have a higher fractional anisotropy in Sandhoff patients compared to Tay-Sachs patients and were observed primarily in the cerebellum (FDR<0.05). No fiber tracts were evaluated to have a higher fractional in Tay-Sachs patients compared to Sandhoff patients (blue).

**Figure 5.**
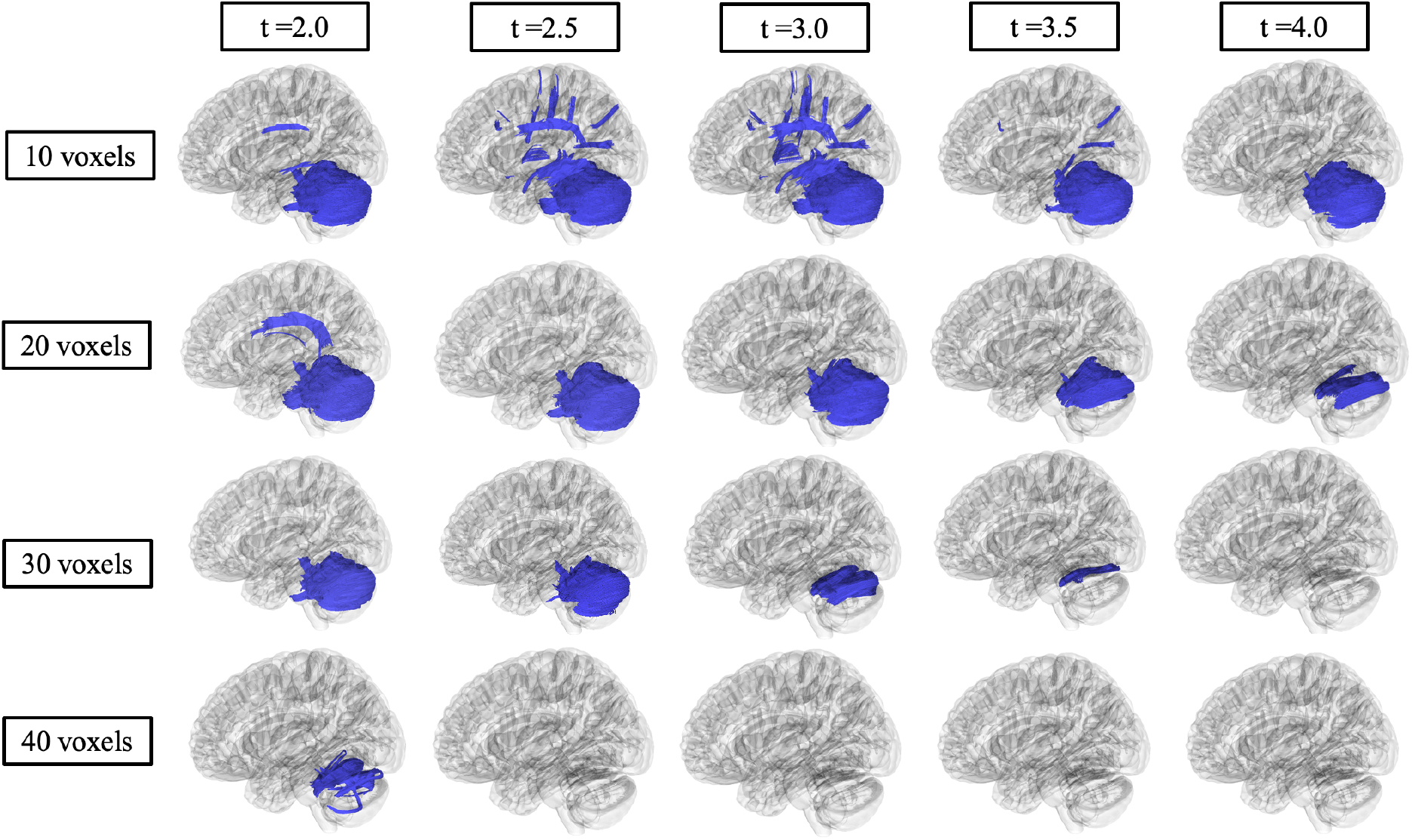
Correlational fiber tractography assessed differences in mean diffusivity in Sandhoff and Tay-Sachs patients at varying length (voxels) and T thresholds. Fiber tracts shown in blue were evaluated to have a higher mean diffusivity in Tay-Sachs patients compared to Sandhoff patients and were observed primarily in the cerebellum (FDR<0.05). No fiber tracts were evaluated to have a higher mean diffusivity in Sandhoff patients compared to Tay-Sachs patients (red).

Correlational fiber tractography results for axial diffusivity (AD), radial diffusivity (RD), and quantitative anisotropy (QA) are shown in Supplement D. AD and RD results were similar to those observed with MD where Tay-Sachs patients were observed to have higher AD and RD in fiber tracts in the cerebellum. QA results were similar to those observed with FA where fiber tracts with higher QA in Sandhoff patients were identified in the cerebellum. QA also highlighted fiber tracts in the brainstem with higher QA in Sandhoff patients, along with a few sparse fiber tracts with higher QA in Tay-Sachs located around the corpus callosum. Higher T-score and length thresholds demonstrated less results and ultimately no results at the highest thresholds for QA, AD and RD.

## Discussion

In this study, we aimed to discern differences in adult onset GM2 gangliosidosis disease subtypes through DWI derived analysis. We further supported the finding of distinct cerebellar pathological differences in Tay-Sachs patients when compared to Sandhoff patients. In 4 out of 7 cerebellar white matter pathways, we found Tay-Sachs patients had reduced FA compared to Sandhoff patients (Table I). In all 7 cerebellar pathways, Tay-Sachs patients also had increased MD, RD, and AD compared to Sandhoff patients (Table II, Supplement Tables C1 and C2). The model free metric QA did not show any significant differences in the 7 cerebellar pathways, the whole brain, or the corpus callosum between Tay-Sachs and Sandhoff patients (Supplement Table C3).

Our correlational fiber tractography results further demonstrate the cerebellar differences between Tay-Sachs and Sandhoff patients. Tay-Sachs patients were observed to have higher MD (Figure 5) in cerebellar fiber tracts compared to Sandhoff. AD and RD (Supplement Figures D1 and D2) correlational fiber tractography results were similar to MD results as both AD and RD were higher in Tay-Sachs patients compared to Sandhoff primarily in cerebellar pathways. Sandhoff patients were observed to have higher FA (Figure 4) in cerebellum tracts compared to Tay-Sachs patients. QA results were similar to our FA result (Supplement Figure D1) where QA was observed to be higher in Sandhoff patients compared to Tay-Sachs patients in cerebellar neuronal pathways. Our QA correlational fiber tractography results also highlight brainstem involvement with lower QA in Tay-Sachs patients when compared with Sandhoff patients which was not observed with any of our DTI metrics (FA/MD/AD/RD).

To determine the sensitivity of our correlational fiber tractography results we tested T-scores from 2.0 to 4.0 and length thresholds from 10 voxels to 40 voxels for all five of our diffusion MRI metrics (QA/FA/MD/AD/RD). As expected, we found significantly fewer correlational tractography results when these thresholds were increased. Higher T-scores are evaluated to have stronger correlations with the study variable, in our case evaluating GM2 gangliosidosis disease subtype. We found significant results in the cerebellum with T-scores between 2.0 and 4.0 for all five of our diffusion MRI metrics as evaluated by correlational tractography when the length threshold was low (10 voxels). When the length threshold was increased, correlational fiber tractography found notably fewer results and ultimately almost no results at the highest length threshold (40 voxels). Previous studies utilizing correlational fiber tractography have utilized T-scores as low as 2.0 and length thresholds as low as 20 voxels to demonstrate correlations with their study variable.^47^ However, correlational fiber tractography T-score and length thresholds in relation to differentiating disease subtypes requires further investigation. Our results also suggest correlational tractography may be an important tool in distinguishing between diseases and disease subtypes with similar neurological clinical presentations or phenotypes.

Limitations of this study need to be considered before these methods and results are used to guide clinical practice or a clinical trial. First, this study is limited by a small sample size (n=16), with four Sandhoff patients and 12 Tay-Sachs patients. Future studies investigating distinctions between Tay-Sachs and Sandhoff patients should include a larger cohort and the addition of neurotypical controls to determine if Sandhoff patients also have subtle cerebellar white matter pathological findings. Similarly, comparison of tractography patterns between Late-onset Tay-Sachs and other more forms of monogenic cerebellar degeneration might be informative and relevant to phenotypic features unique to Tay-Sacks disease. Secondly, the use of FA, and particularly both AD and RD as biomarkers for white matter disease have been met with skeptisism.^48^ Voxels with crossing white matter fibers and complex fiber geometry can lead to incorrect interpretations of these metrics when taken in isolation.^48^ The present study addresses this issue by first taking into consideration MD which is more robust than FA, AD, and RD. Secondly, these metrics were not interpreted in isolation, and they supported the same conclusion of distinct cerebellar pathology in Tay-Sachs patients when compared to Sandhoff patients. Lastly, the present study focuses solely on diffusion tensor imaging (DTI), future studies should investigate neurite orientation dispersion and density imaging (NODDI),^49^ metabolic activity diffusion imaging (MADI),^50^ volumetric analysis of T1-weighted MRI data, and magnetic resonance spectroscopy to define the full neuroimaging phenotype of late-onset GM2 gangliosidosis patients.

## Conclusion

In this study, we aimed to describe differences in diffusion MRI metrics between adult Sandhoff and adult Tay-Sachs patients. Our DTI analysis found differences between Tay-Sachs and Sandhoff patients primarily in cerebellar pathways in FA, MD, RD, and AD, suggesting altered white matter cerebellar pathology in Tay-Sachs patients. To our knowledge, this is the first study using correlational tractography in a lysosomal storage disorder to demonstrates differences in disease subtypes. FA decreases and MD, AD, and RD increases were observed in Tay-Sachs patients when compared to Sandhoff patients in cerebellum fiber tracts which further supports the result of distinct cerebellar pathology in Tay-Sachs patients. Cerebellar symptoms are more common in Tay-Sachs patients, which could be caused by the cerebellar white matter pathology observed in this study.

## Data Availability

The data described in this manuscript are available from the corresponding author upon reasonable request.

## Acknowledgements

We thank the participants and their families for the generosity of their time and efforts. We are also grateful to many staff members and care providers who contributed their expertise over the years. DWI preprocessing in this work utilized the computational resources of the Biowulf Linux cluster at the National Institutes of Health (http://hpc.nih.gov).

## Author Contributions

Conceptualization: CJL, CT, CJT; Data Curation: PD, JMJ, CJT, MTA; Funding Acquisition: CJT; Methodology: CJL, SIC, CT, MTA, CJT; Visualization: CJL, SIC, CT, MTA, CJT; Writing-original draft: CJL, CJT; Writing-reviewing & editing; CJL, SIC, PD, JMJ, CT, MTA, CJT

## Ethics Declaration

The NIH Institutional Review Board approved this protocol (02-HG-0107). Informed consent was completed with parents or legal guardians of the patients. All participants were assessed for their ability to provide assent; none were deemed capable.

## Abbreviations

AD: Axial Diffusivity
DTI: Diffusion Tensor Imaging
DWI: Diffusion Weighted Imaging FA Fractional Anisotropy
LOSD: Late-onset Sandhoff Disease
LOTS: Late-onset Tay-Sachs
MD: Mean Diffusivity
MRI: Magnetic Resonance Imaging
MRS: Magnetic Resonance Spectroscopy
QA: Quantitative Anisotropy
RD: Radial Diffusivity

## Supplementary Methods

### Supplement A: Natural History Study Participant Characteristics

**Table A1.**
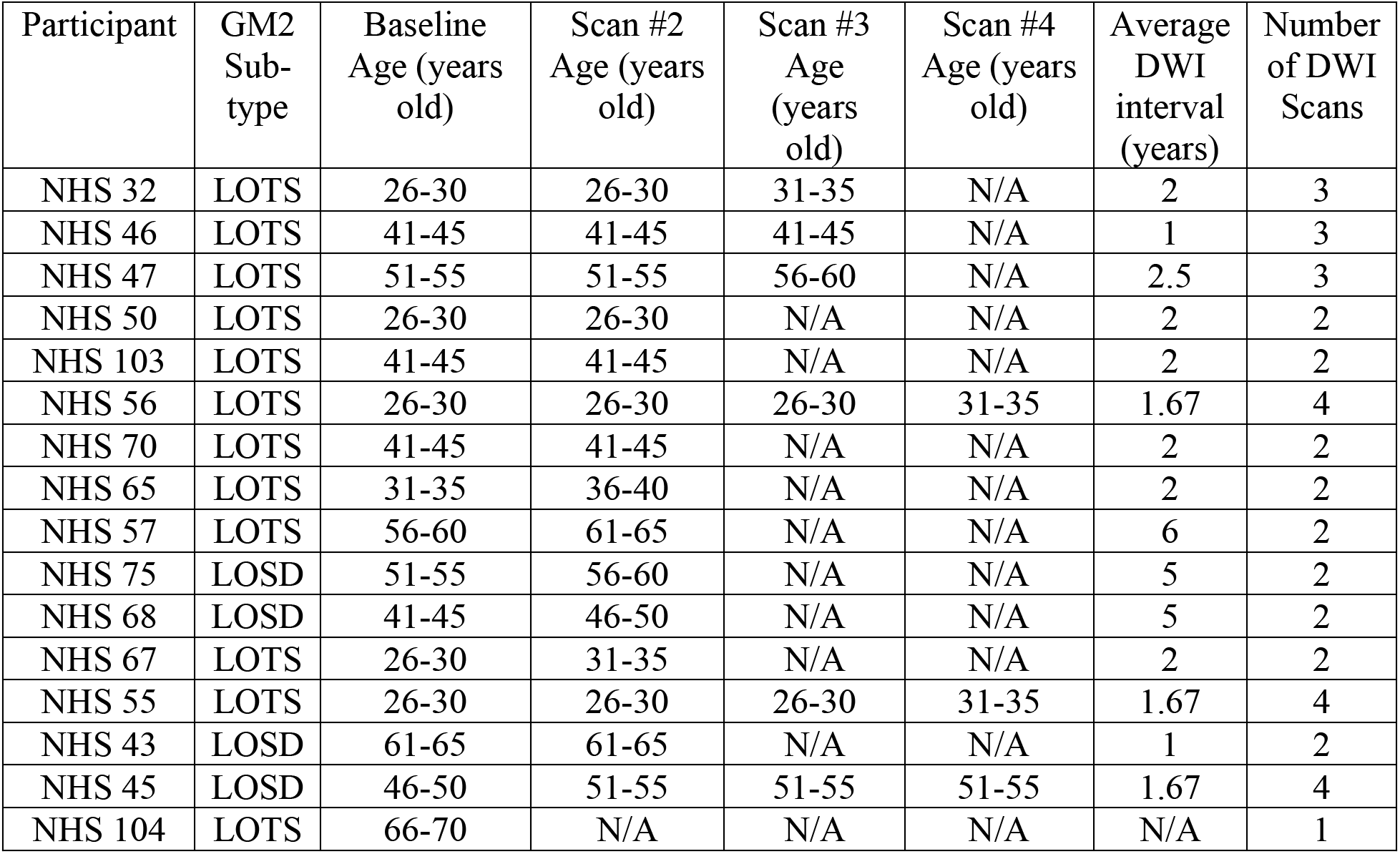
Natural History Study DWI Cohort (n = 16), specific ages redacted per MedArXiv requirements.

## Supplementary Results

### Supplement B: Diffusion Tensor Imaging Analysis

#### Supplement Figures

**Figure B1.**
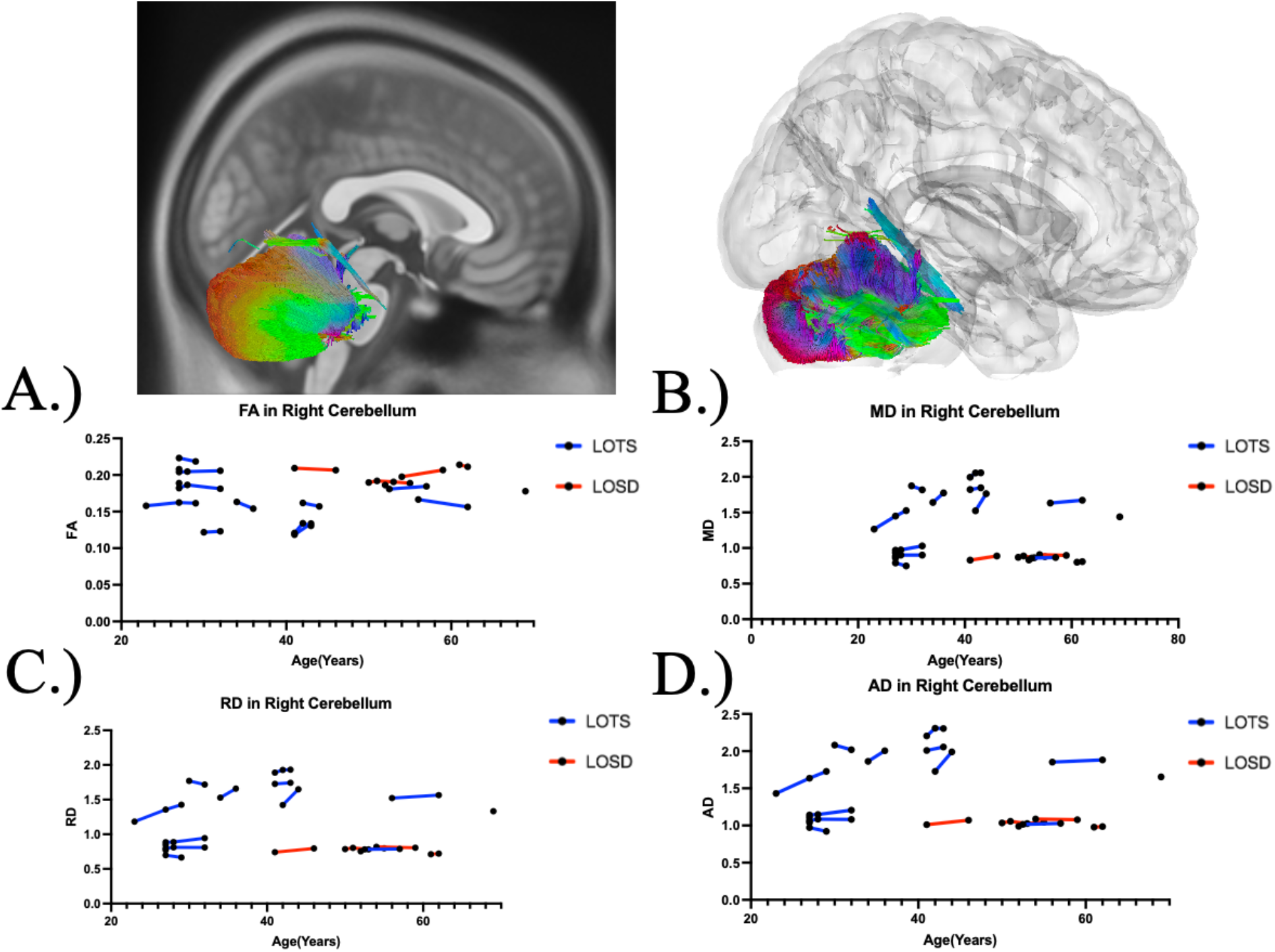
Atlas based fiber tractography of the right cerebellum demonstrating age related effects on A.) fractional anisotropy B.) mean diffusivity between C.) radial diffusivity D.) axial diffusivity between Tay-Sachs patients (blue) and Sandhoff patients (red). Tay-Sachs patients demonstrated decreased FA (*χ*^2^(1) = 5.50, *p =* 0.019) and increased MD (*χ*^2^(1) = 8.56, *p =* 0.0034), RD (*χ*^2^(1) = 8.55, *p =* 0.0035), and AD (*χ*^2^(1) = 8.57, *p =* 0.0034) compared to Sandhoff patients in fiber tracts in the right cerebellum when age was accounted for.

**Fig B2.**
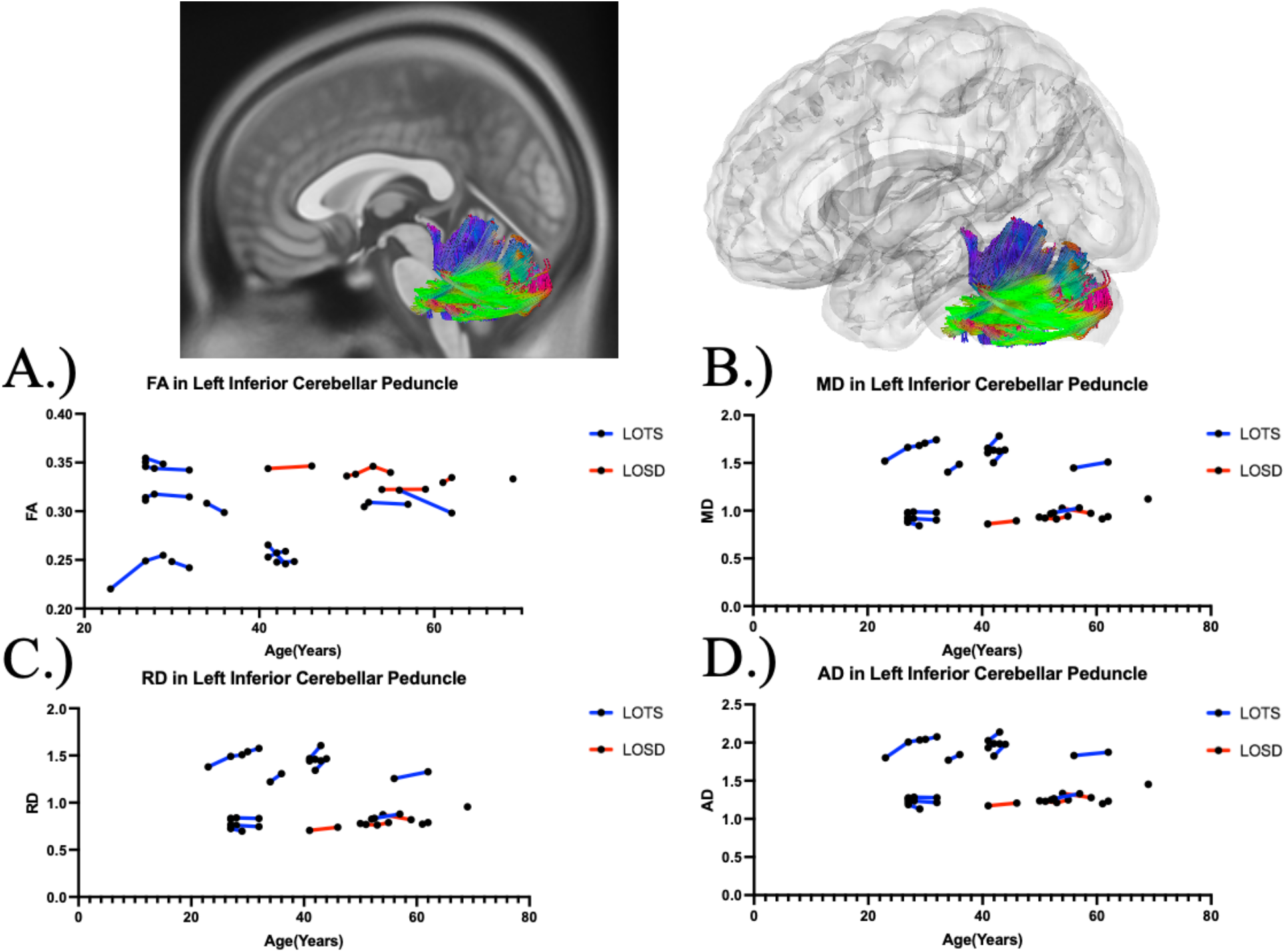
Atlas based fiber tractography of the left inferior cerebellar peduncle demonstrating age related effects on A.) fractional anisotropy B.) mean diffusivity between C.) radial diffusivity D.) axial diffusivity between Tay-Sachs patients (blue) and Sandhoff patients (red). Tay-Sachs patients demonstrated no difference in FA (*χ*^2^(1) = 3.10, *p =* 0.078) and increased MD (*χ*^2^(1) = 7.02, *p =* 0.0081), RD (*χ*^2^(1) = 6.85, *p =* 0.0089), and AD (*χ*^2^(1) = 7.26, *p =* 0.0070) compared to Sandhoff patients in fiber tracts in the left inferior cerebellar peduncle when age was accounted for.

**Fig B3.**
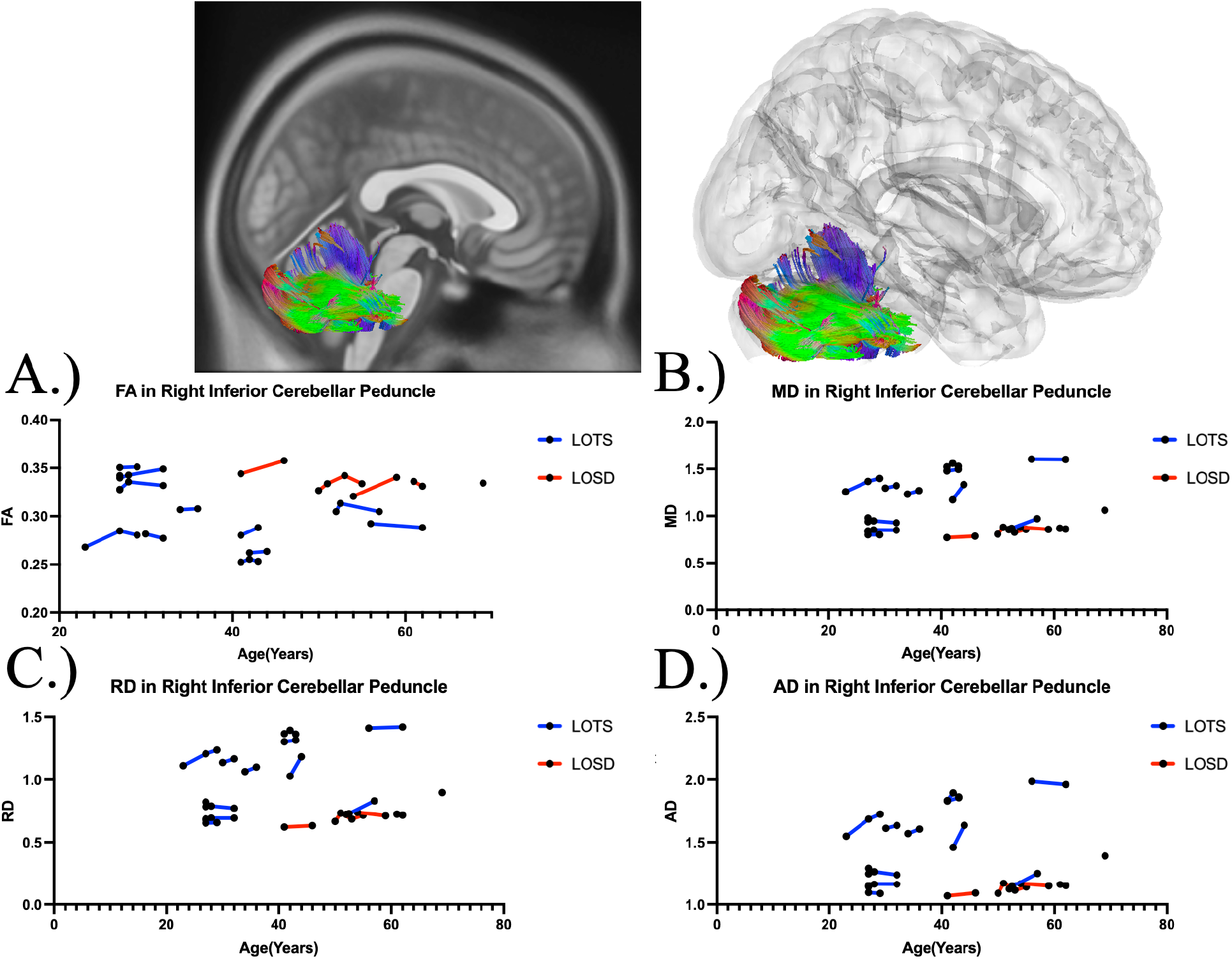
Atlas Based Fiber Tractography of the right inferior cerebellar peduncle demonstrating age related effects on A.) fractional anisotropy B.) mean diffusivity between Tay-Sachs patients (blue) and Sandhoff patients (red). Tay-Sachs patients demonstrated no difference in FA (*χ*^2^(1) = 1.89, *p =* 0.17) and increased MD (*χ*^2^(1) = 8.55, *p =* 0.0035), RD (*χ*^2^(1) = 8.39, *p =* 0.0038), and AD (*χ*^2^(1) = 8.79, *p =* 0.0030) compared to Sandhoff patients in fiber tracts in the right inferior cerebellar peduncle when age was accounted for.

**Fig B4.**
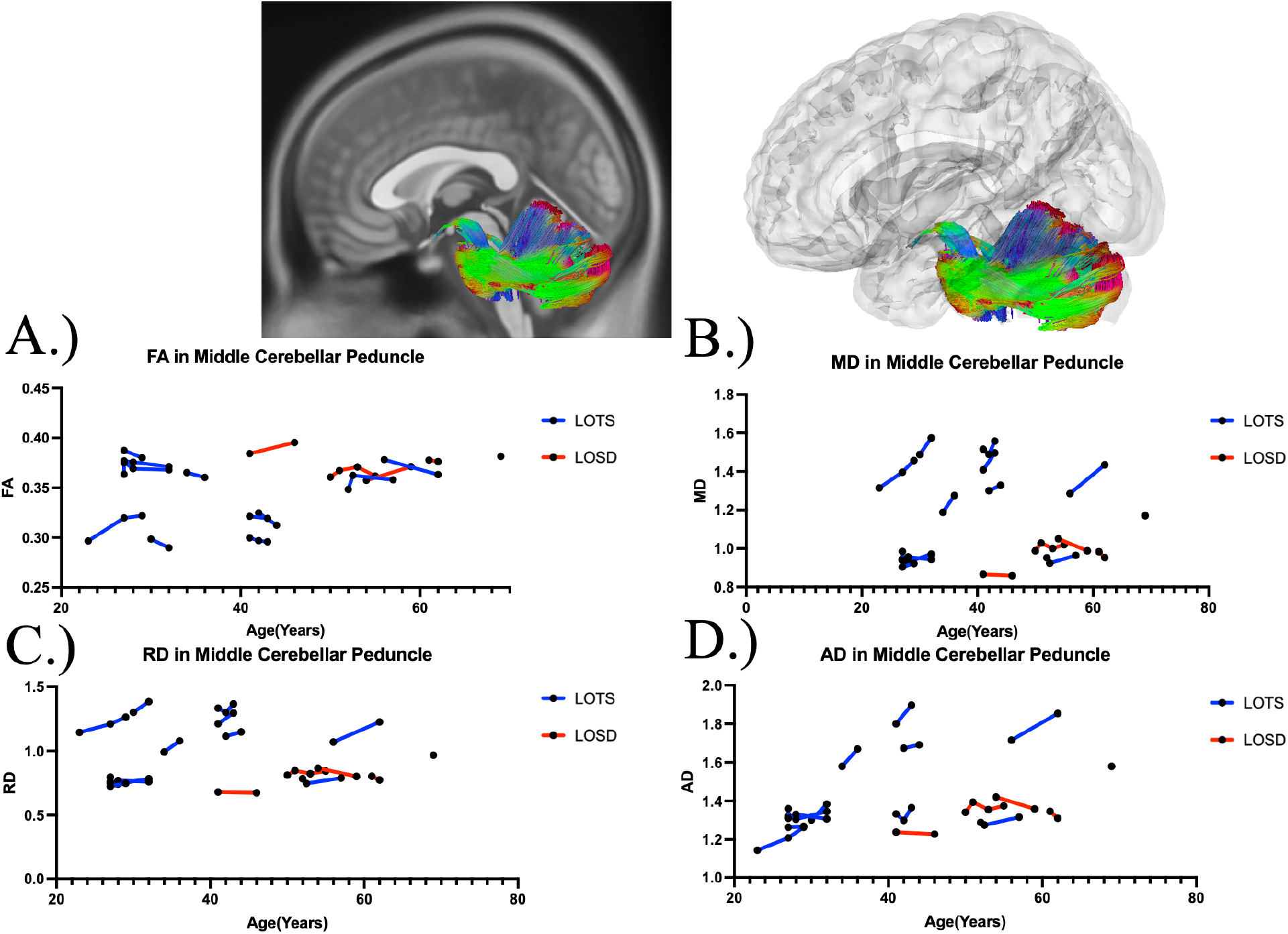
Atlas Based Fiber Tractography of the middle cerebellar peduncle demonstrating age related effects on A.) fractional anisotropy B.) mean diffusivity between Tay-Sachs patients (blue) and Sandhoff patients (red). Tay-Sachs patients demonstrated no difference in FA (*χ*^2^(1) = 1.65, *p =* 0.20) and increased MD (*χ*^2^(1) = 6.81, *p =* 0.0091), RD (*χ*^2^(1) = 6.55, *p =* 0.010), and AD (*χ*^2^(1) = 7.22, *p =* 0.0072) compared to Sandhoff patients in fiber tracts in the middle cerebellar peduncle when age was accounted for.

**Fig B5.**
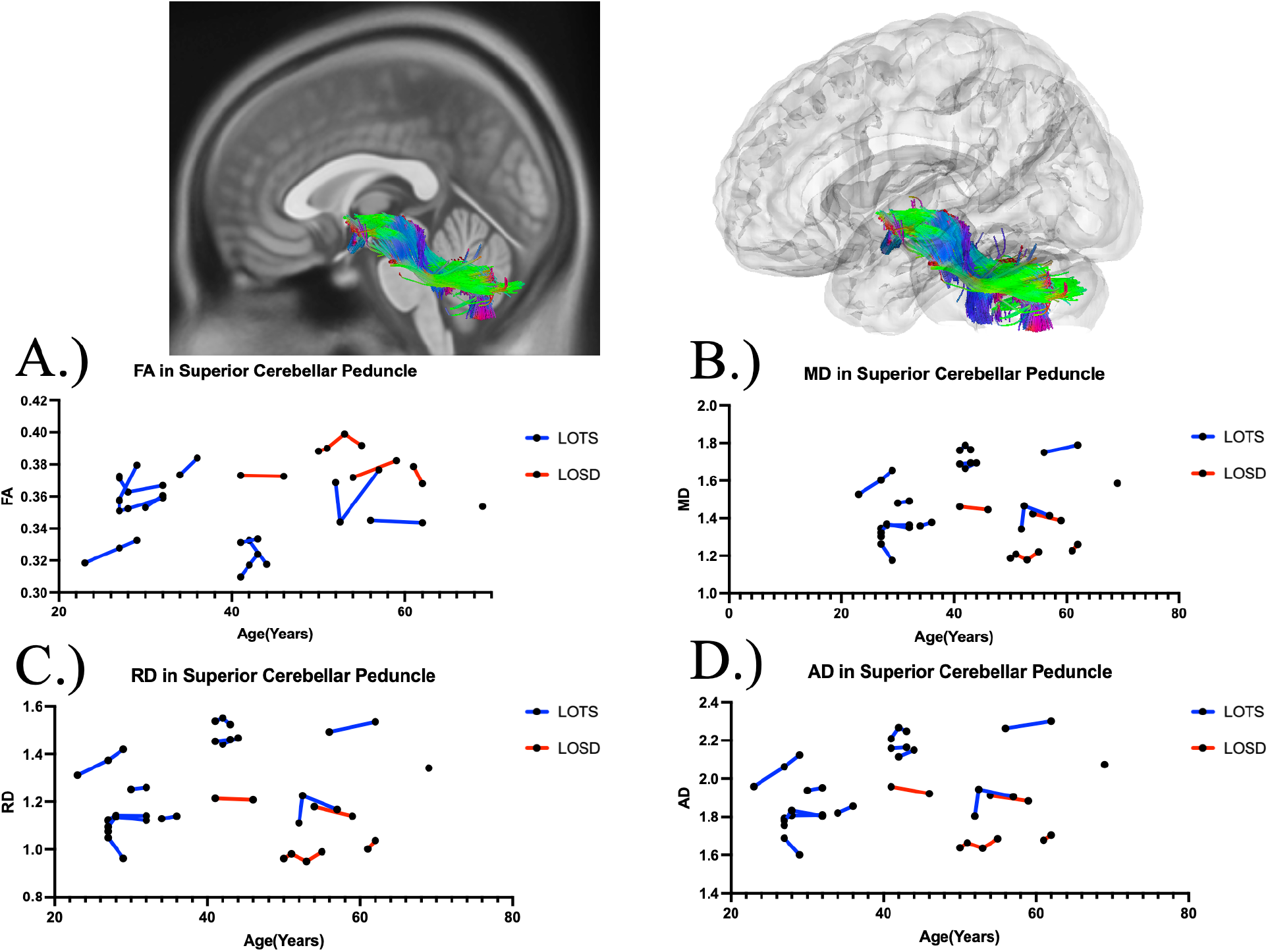
Atlas Based Fiber Tractography of the superior cerebellar peduncle demonstrating age related effects on A.) fractional anisotropy B.) mean diffusivity between Tay-Sachs patients (blue) and Sandhoff patients (red). Tay-Sachs patients demonstrated lower FA (*χ*^2^(1) = 4.80, *p =* 0.028) and increased MD (*χ*^2^(1) = 7.19, *p =* 0.0073), RD (*χ*^2^(1) = 7.18, *p =* 0.0074), and AD (*χ*^2^(1) = 7.09, *p =* 0.0078) compared to Sandhoff patients in fiber tracts in the superior cerebellar peduncle when age was accounted for.

**Fig B6.**
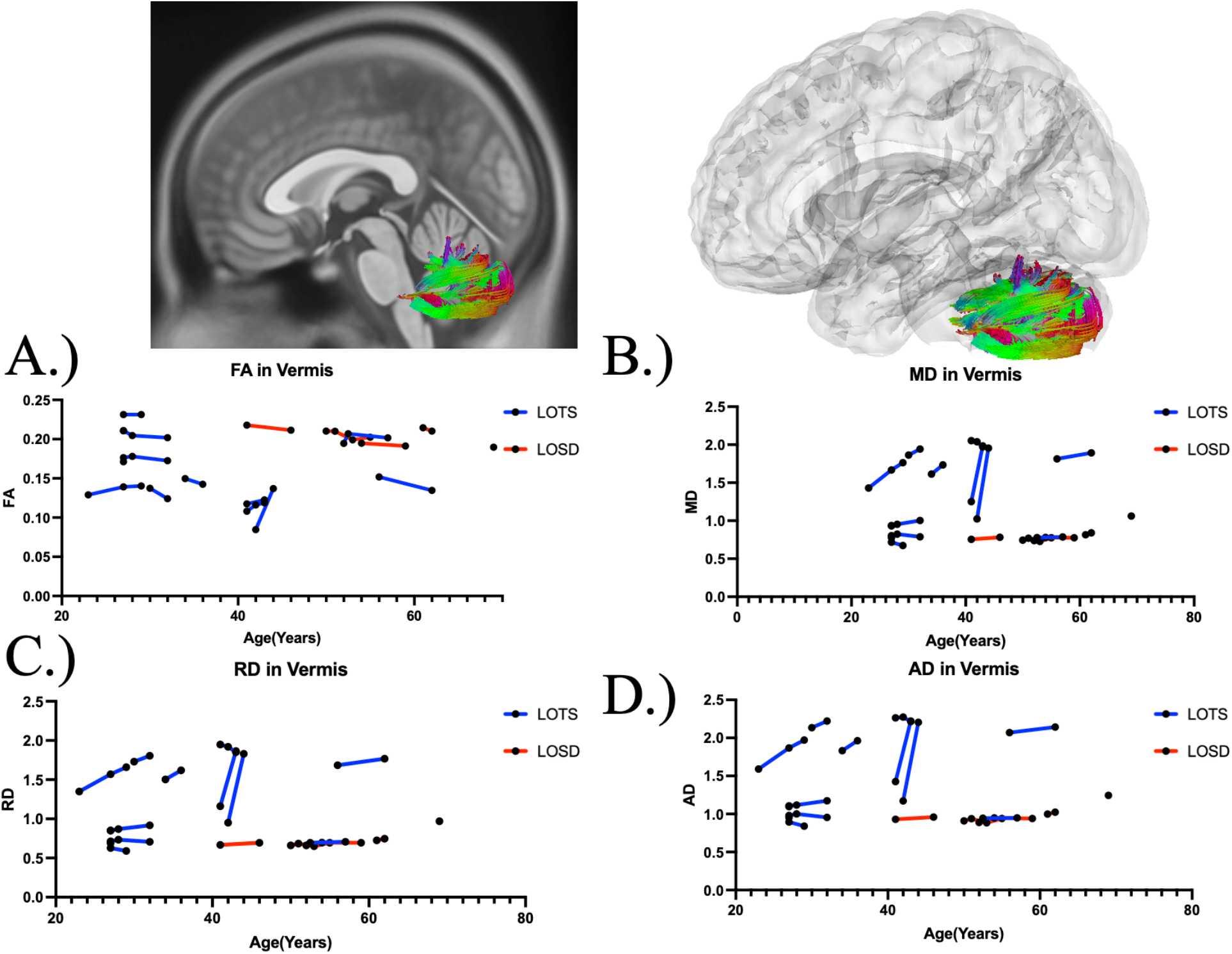
Atlas Based Fiber Tractography of the cerebellar vermis demonstrating age related effects on A.) fractional anisotropy B.) mean diffusivity between Tay-Sachs patients (blue) and Sandhoff patients (red). Tay-Sachs patients demonstrated lower FA (*χ*^2^(1) = 5.05, *p =* 0.025) and increased MD (*χ*^2^(1) = 6.62, *p =* 0.010), RD (*χ*^2^(1) = 6.70, *p =* 0.0096), and AD (*χ*^2^(1) = 6.47, *p =* 0.011) compared to Sandhoff patients in fiber tracts in the cerebellar vermis when age was accounted for.

**Fig B7.**
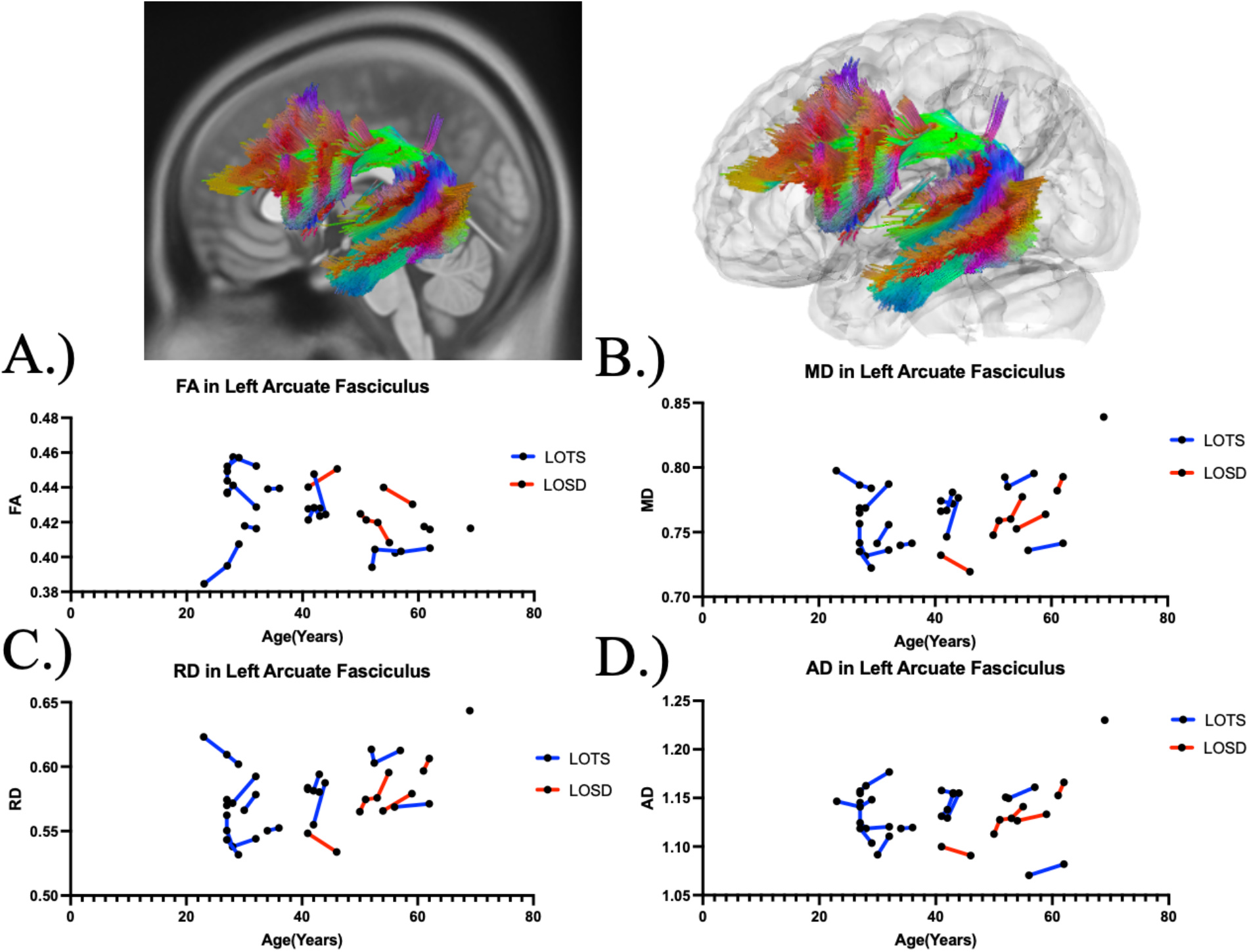
Atlas Based Fiber Tractography of the left arcuate fasciculus demonstrating age related effects on A.) fractional anisotropy B.) mean diffusivity between Tay-Sachs patients (blue) and Sandhoff patients (red). There was no statistical difference between Tay-Sachs and Sandhoff patients in FA (*χ*^2^(1) = 0.92, *p =* 0.34), MD (*χ*^2^(1) = 2.65, *p =* 0.10), RD (*χ*^2^(1) = 2.66, *p =* 0.10), or AD (*χ*^2^(1) = 2.02, *p =* 0.16) in fiber tracts in the left arcuate fasciculus.

**Fig B8.**
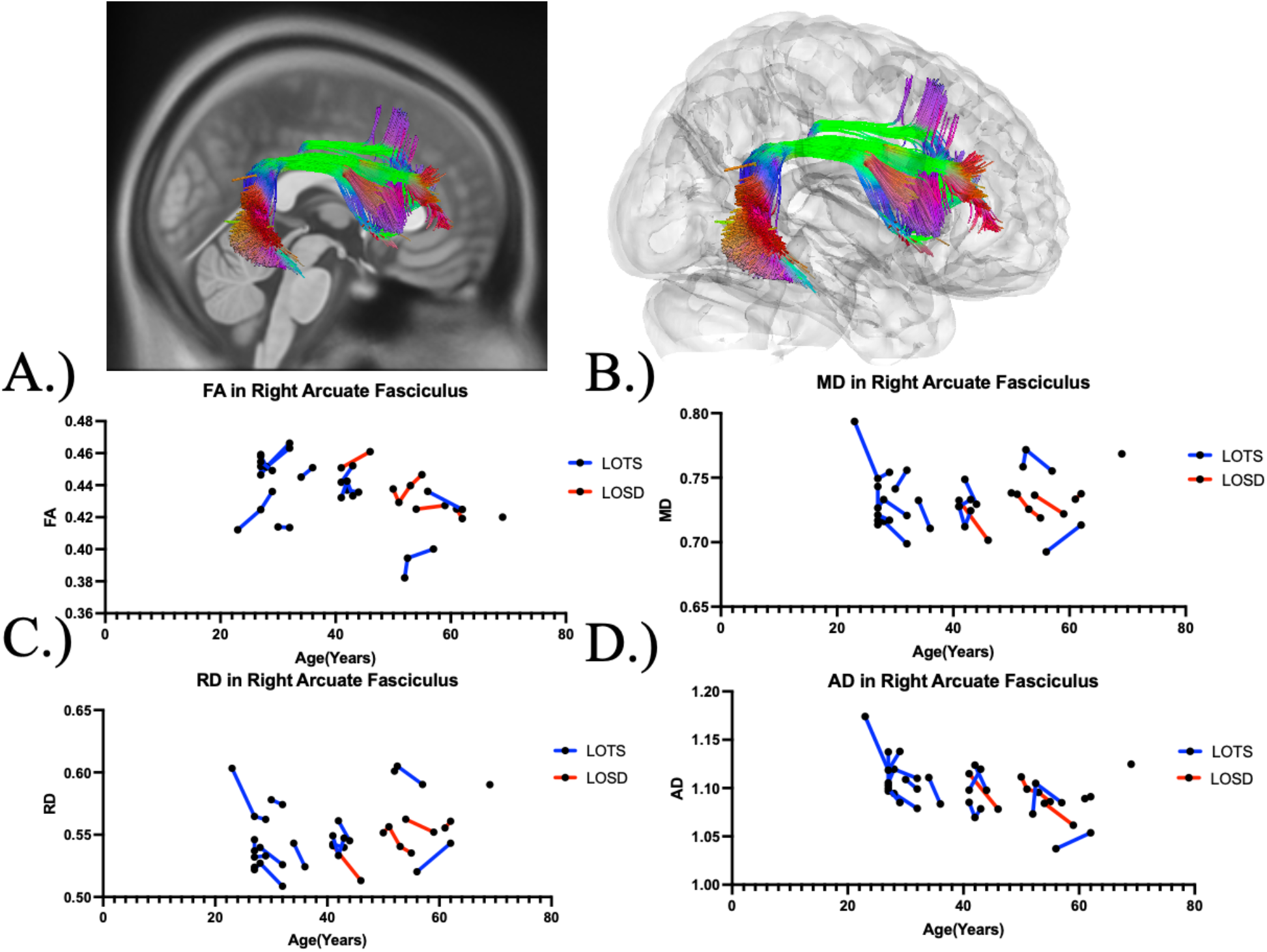
Atlas Based Fiber Tractography of the right arcuate fasciculus demonstrating age related effects on A.) fractional anisotropy B.) mean diffusivity between Tay-Sachs patients (blue) and Sandhoff patients (red). There was no statistical difference between Tay-Sachs and Sandhoff patients in FA (*χ*^2^(1) = 0.044, *p =* 0.834), MD (*χ*^2^(1) = 0.12, *p =* 0.73), RD (*χ*^2^(1) = 0.14, *p =* 0.71), or AD (*χ*^2^(1) = 0.079, *p =* 0.78) in fiber tracts in the right arcuate fasciculus.

**Figure B9.**
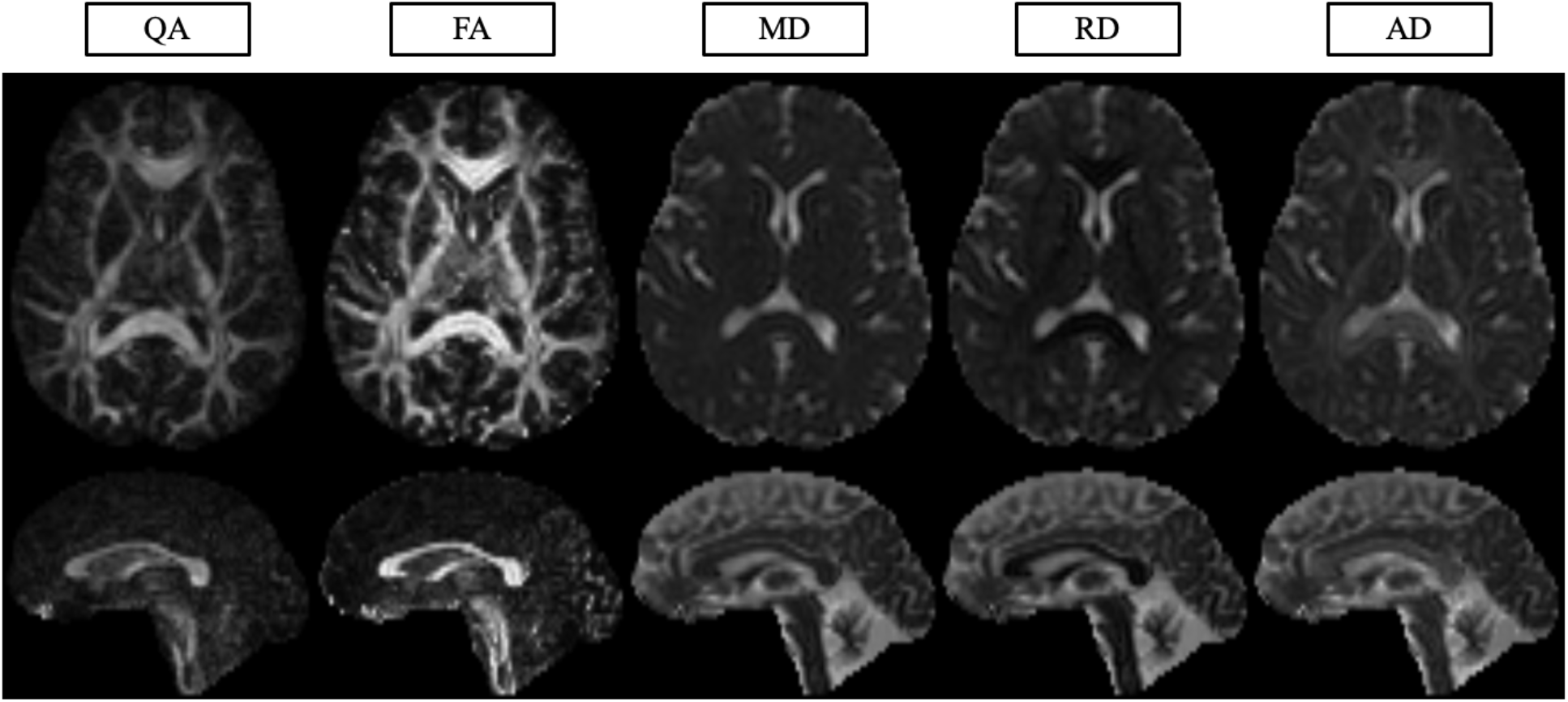
Quantitative Anisotropy (QA), Fractional Anisotropy (FA), Mean Diffusivity (MD), Radial Diffusivity (RD), and Axial Diffusivity (AD) imaging.

### Supplement C. Diffusion Tensor Imaging Analysis of RD, AD, and QA

**Table C1.**
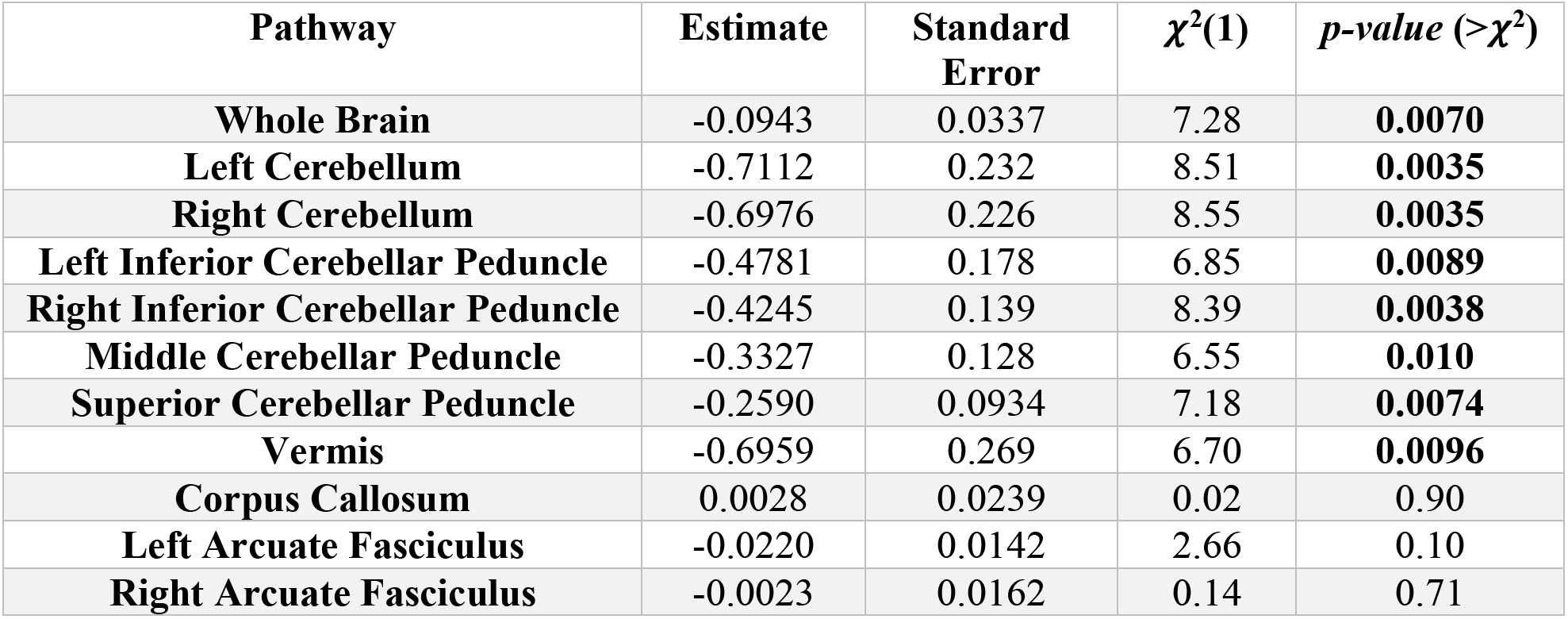
Diffusion Tensor Imaging Results of RD in Atlas Fiber Tractography Pathways evaluating differences between Tay-Sachs and Sandhoff Patients.

| Pathway | Estimate | Standard Error | $\chi^2(1)$ | $p$-value ($>\chi^2$) |
| --- | --- | --- | --- | --- |
| Whole Brain | -0.0943 | 0.0337 | 7.28 | **0.0070** |
| Left Cerebellum | -0.7112 | 0.232 | 8.51 | **0.0035** |
| Right Cerebellum | -0.6976 | 0.226 | 8.55 | **0.0035** |
| Left Inferior Cerebellar Peduncle | -0.4781 | 0.178 | 6.85 | **0.0089** |
| Right Inferior Cerebellar Peduncle | -0.4245 | 0.139 | 8.39 | **0.0038** |
| Middle Cerebellar Peduncle | -0.3327 | 0.128 | 6.55 | **0.010** |
| Superior Cerebellar Peduncle | -0.2590 | 0.0934 | 7.18 | **0.0074** |
| Vermis | -0.6959 | 0.269 | 6.70 | **0.0096** |
| Corpus Callosum | 0.0028 | 0.0239 | 0.02 | 0.90 |
| Left Arcuate Fasciculus | -0.0220 | 0.0142 | 2.66 | 0.10 |
| Right Arcuate Fasciculus | -0.0023 | 0.0162 | 0.14 | 0.71 |

**Table C2.**
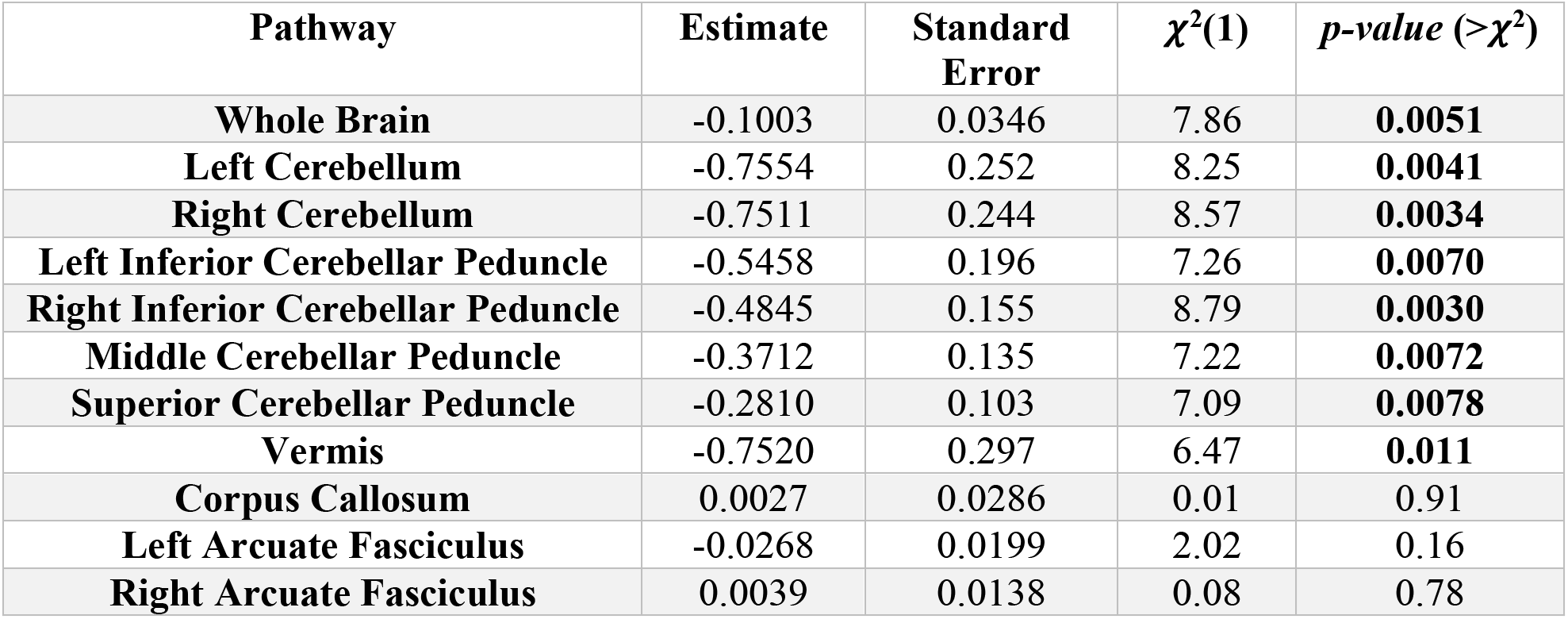
Diffusion Tensor Imaging Results of AD in Atlas Fiber Tractography Pathways evaluating differences between Tay-Sachs and Sandhoff Patients.

| Pathway | Estimate | Standard Error | $\chi^2(1)$ | $p$-value ($>\chi^2$) |
| --- | --- | --- | --- | --- |
| Whole Brain | -0.1003 | 0.0346 | 7.86 | **0.0051** |
| Left Cerebellum | -0.7554 | 0.252 | 8.25 | **0.0041** |
| Right Cerebellum | -0.7511 | 0.244 | 8.57 | **0.0034** |
| Left Inferior Cerebellar Peduncle | -0.5458 | 0.196 | 7.26 | **0.0070** |
| Right Inferior Cerebellar Peduncle | -0.4845 | 0.155 | 8.79 | **0.0030** |
| Middle Cerebellar Peduncle | -0.3712 | 0.135 | 7.22 | **0.0072** |
| Superior Cerebellar Peduncle | -0.2810 | 0.103 | 7.09 | **0.0078** |
| Vermis | -0.7520 | 0.297 | 6.47 | **0.011** |
| Corpus Callosum | 0.0027 | 0.0286 | 0.01 | 0.91 |
| Left Arcuate Fasciculus | -0.0268 | 0.0199 | 2.02 | 0.16 |
| Right Arcuate Fasciculus | 0.0039 | 0.0138 | 0.08 | 0.78 |

**Table C3.** Diffusion Tensor Imaging Results of QA in Atlas Fiber Tractography Pathways evaluating differences between Tay-Sachs and Sandhoff Patients.

| **Pathway** | **Estimate** | **Standard Error** | **$\chi^2(1)$** | ***p-value* ($>\chi^2$)** |
| --- | --- | --- | --- | --- |
| **Whole Brain** | -0.0084 | 0.0109 | 0.68 | 0.41 |
| **Left Cerebellum** | 0.0337 | 0.0185 | 3.57 | 0.06 |
| **Right Cerebellum** | 0.0193 | 0.0168 | 1.53 | 0.22 |
| **Left Inferior Cerebellar Peduncle** | 0.0208 | 0.0201 | 1.15 | 0.28 |
| **Right Inferior Cerebellar Peduncle** | 0.0024 | 0.0169 | 0.02 | 0.90 |
| **Middle Cerebellar Peduncle** | 0.0240 | 0.0206 | 1.47 | 0.22 |
| **Superior Cerebellar Peduncle** | 0.0065 | 0.0155 | 0.16 | 0.69 |
| **Vermis** | 0.0310 | 0.0191 | 2.89 | 0.09 |
| **Corpus Callosum** | -0.0214 | 0.0137 | 2.65 | 0.10 |
| **Left Arcuate Fasciculus** | -0.0107 | 0.0132 | 0.73 | 0.39 |
| **Right Arcuate Fasciculus** | -0.0211 | 0.0143 | 2.39 | 0.12 |

### Supplement D: Correlational Fiber Tractography Analysis, Supplement Figures

**Figure D1.**
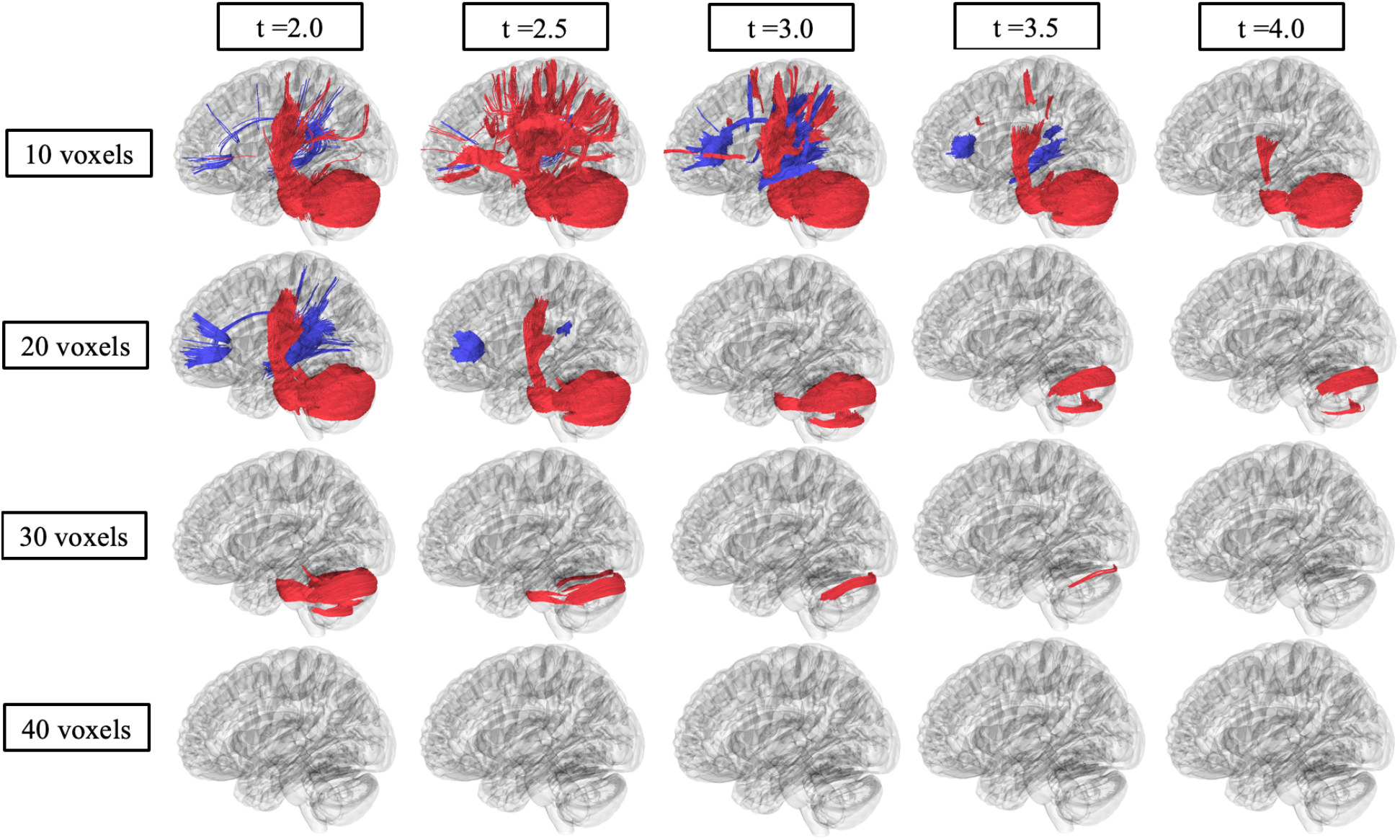
Correlational fiber tractography assessed differences in quantitative anisotropy (QA) in Sandhoff and Tay-Sachs patients at varying length (voxels) and T thresholds. Fiber tracts shown in red were evaluated to have a higher quantitative anisotropy in Sandhoff patients compared to Tay-Sachs patients and were observed primarily in the cerebellum (FDR<0.05). Fiber tracts shown in blue were evaluated to have a higher quantitative anisotropy in Tay-Sachs patients compared to Sandhoff patients.

**Figure D2.**
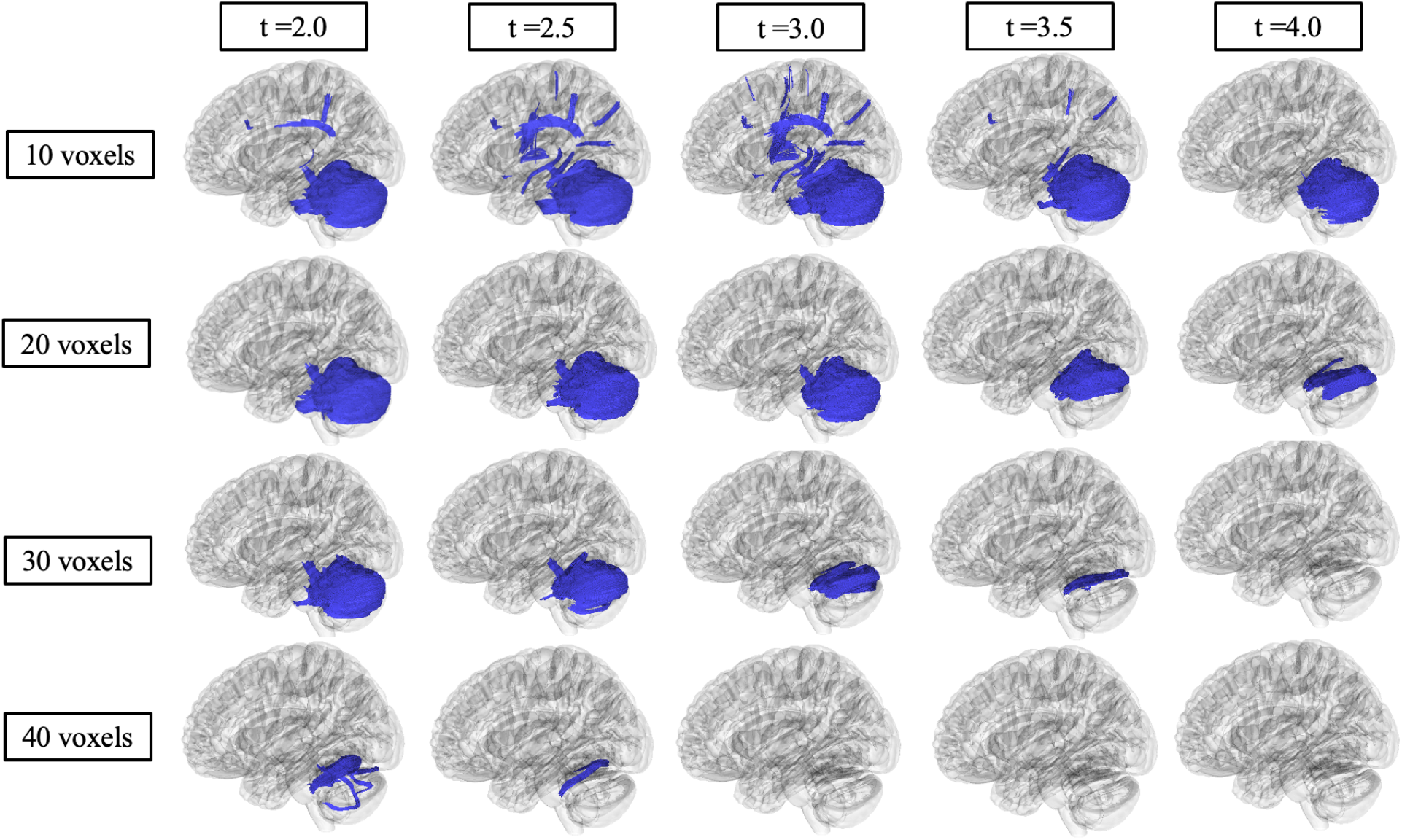
Correlational fiber tractography assessed differences in radial diffusivity (RD) in Sandhoff and Tay-Sachs patients at varying length (voxels) and T thresholds. Fiber tracts shown in blue were evaluated to have a higher radial diffusivity in Tay-Sachs patients compared to Sandhoff patients and were observed primarily in the cerebellum (FDR<0.05). No fiber tracts were evaluated to have a higher radial diffusivity in Sandhoff patients compared to Tay-Sachs patients (red).

**Figure D3.**
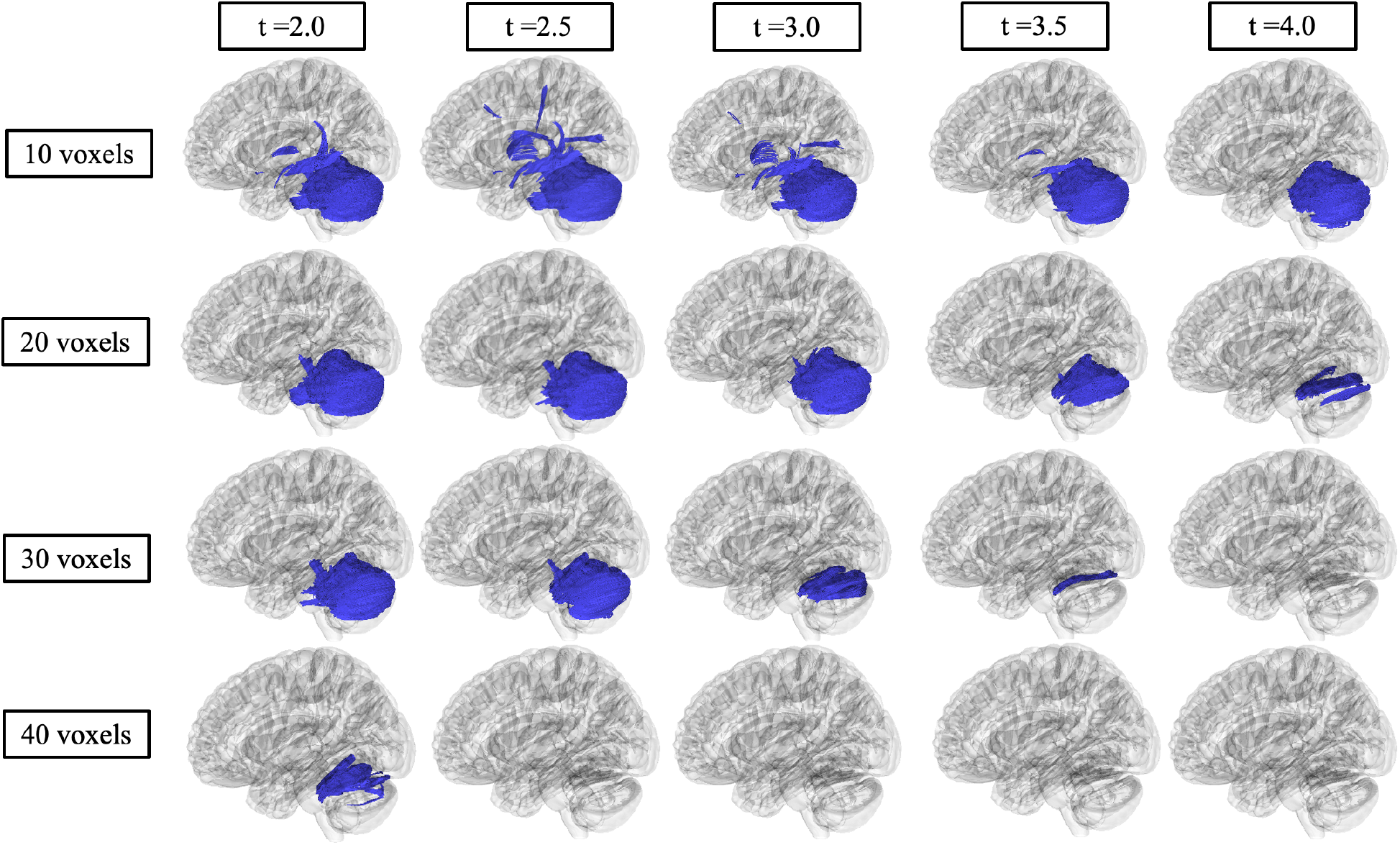
Correlational fiber tractography assessed differences in axial diffusivity (AD) in Sandhoff and Tay-Sachs patients at varying length (voxels) and T thresholds. Fiber tracts shown in blue were evaluated to have a higher axial diffusivity in Tay-Sachs patients compared to Sandhoff patients and were observed primarily in the cerebellum (FDR<0.05). No fiber tracts were evaluated to have a higher axial diffusivity in Sandhoff patients compared to Tay-Sachs patients (red).

